# INCREASED PREVALENCE OF AUTISM IN THE CHILDREN OF FIBROMYALGIA PATIENTS RELATIVE TO OTHER TYPES OF CHRONIC PAIN

**DOI:** 10.1101/2025.10.20.25336737

**Authors:** A. Hirst, R. Mountford, R. Berwick, P. Christiansen, S. Sandin, W. Yin, D A. Andersson, F. Happé, N. Fallon, A. Goebel

## Abstract

Fibromyalgia syndrome (FMS) is a chronic primary pain condition characterised by widespread pain and hypersensitivities, and symptoms of fatigue, poor sleep and impaired cognition. The current studies were prompted by clinical observations made at a tertiary pain centre, where patients with severe FMS frequently reported having biological children on the autism spectrum. Using multimodal data collection techniques, including a clinical service evaluation (N = 589), an exploratory phenotyping study (N= 59), an online patient survey (N= 710), and a clinical prospective investigation (N = 66), the present study aimed to investigate links that may point to a connection between maternal FMS and autistic offspring. For the clinical and online data collection, the studies used comparator patients with complex regional pain syndrome (CRPS) and a mix of “other chronic pain conditions”. Maternal factors of interest, including a familial history of neurodivergence and psychological variables, were considered in the analysis. Maternal FMS was consistently found to be associated with an increased likelihood of children with an autism diagnosis, even after accounting for established maternal factors associated with autism. Meeting and advocating for the support needs of autistic children may require enhanced care, which can lead to greater demands on patients already suffering from FMS or CRPS. The causes for the observed association between maternal FMS and autistic children require further investigation.

## Introduction

Fibromyalgia syndrome (FMS) is a chronic primary pain condition characterised by symptoms of widespread musculoskeletal pain, fatigue, stiffness, and “brain fog” [11; 35]. It is frequently associated with other somatic (e.g., irritable bowel syndrome) and psychological (e.g., anxiety, depression, PTSD) comorbidities [20]. The estimated global prevalence of FMS is commonly reported at 2-3% [36], with UK estimates reaching up to 5.4% [17]. Sixty-90% of cases are women [12; 46]. The pathogenesis of FMS is currently unknown, however, accumulating evidence has suggested that immune system dysfunction could underlie characteristic symptoms [19; 24; 32]. Several features of FMS are suggestive of autoimmunity, including the female predominance and mid-age onset [12], the notable prevalence of autoimmune disease comorbidity [2; 18], gene expression profiles which are typical of autoimmune disease [14], the relationship between the onset of FMS symptoms and trauma and infection [47], and the presence of autoreactive antibodies [5; 19].

Autism spectrum disorder (hereafter referred to as autism) is a neurodevelopmental condition characterised by social interaction and communication difficulties along with a range of restricted and/or repetitive patterns of behaviour, interests, or activities [3]. Moreover, autism presents with atypical sensorimotor function and often co-occurs with chronic pain [7; 45]. The estimated prevalence of autism in the general UK population is between 1% and 3% [8; 33], however, it is known to be underdiagnosed in older adults and, historically, in women. Although the exact aetiology of autism is not yet known, a complex interplay involving genetic and environmental factors is proposed [27]. Immune system dysregulation has been suggested as a non-genetic contributing factor and has been described in autistic people and their family members [15; 40]. Previous research has also implicated the prenatal immune environment as a factor in autism development [25; 37; 38], and several studies have demonstrated an increased likelihood of autistic children of mothers, but not fathers, with autoimmune diseases [10; 13; 48].

In clinical observations made by a consultant in pain medicine at a tertiary pain centre (AG), it was noted that patients with severe FMS frequently reported having biological children on the autism spectrum. Caring and advocating for an autistic child can place additional demands on the parent [34], which may be particularly challenging for people suffering from severe chronic pain. It was thought, therefore, that understanding any such association between FMS and autistic offspring would be relevant for practitioners in this field, enabling them to gather information pertinent to gauge support requirements.

## Methods

The study used a design in which data from multiple, independent sources were compiled to investigate a potential link between maternal FMS and children with an autism diagnosis. This comprised data from a clinical pain centre service evaluation (Study A1), a previously established research study (Study A2), and two newly conducted online surveys – one using a wider geographical region in the UK (Study B) and one using a subset of patients included in the clinical service evaluation (Study C).

### A1) Clinical Service Evaluation

#### Participants

Following initial observations about autistic children of patients with FMS at a tertiary pain centre, we systematically documented the frequency of autism diagnoses in the offspring of patients with FMS and other chronic pain conditions at the same institution (the Walton Centre, a UK neuro-care centre).

Chronic pain patients were included in a service evaluation if they had attended their appointment as a new patient at the tertiary care outpatient pain clinic with a consultant in pain medicine (AG) between 26^th^ October 2020 and 10^th^ May 2023. All patients for whom a clinic letter was dictated during this period were included (N=589). As part of their routine consultation, the consultant had asked new patients clinical questions, which for the majority included a question about their children’s health. The consultant did not specifically enquire about neurodivergent diagnoses in children in any of the available clinic letters, and no questionnaires were used. The service evaluation was approved by the Walton Centre NHS Foundation Trust (NS451).

#### Diagnostic categorisation

The recruiting centre provides health care in matters of pain from the north-west region of England and is also a supra-regional referral centre for persistent Complex Regional Pain Syndrome (CRPS) and other uncommon, complex chronic pain conditions. Patients were categorised by their chronic pain condition: FMS, persistent CRPS, and other chronic pain conditions. In the “other chronic pain condition” group, a majority suffered from chronic widespread pain or back and leg pain conditions. A smaller group suffered from neuropathic pain conditions or other chronic pain (for details, see Supplementary Table 1; Appendix A).

Patients who reported their children as having an autism diagnosis were classified as cases in the analysis. Patients who reported their child as having *“suspected*” or “*probable”* autism were not classified as a case of offspring autism in the analysis.

#### Statistical analysis

Penalised likelihood logistic regression analysis was conducted to test the association between FMS and CRPS, compared to other chronic pain conditions, and the likelihood of offspring with autism diagnoses. Firth’s penalised logistic regression is a method for modelling binary outcomes, used to mitigate issues that traditional logistic regression models may have when facing small sample sizes, rare events, and separation [43]. The categorical variable “chronic pain condition” had 3 levels (FMS, CRPS, and other chronic pain). Other chronic pain was chosen as the baseline category to observe any change in log-odds for FMS and CRPS, which are particularly severe chronic primary pain conditions, previously implicated as having an autoimmune mechanism [19; 23]. At present, no such evidence has emerged for the other chronic pain group. Only mothers with confirmed biological children were included in the analysis. Odds ratios (OR) with 95% confidence intervals (CIs) were calculated, corresponding to a two-sided test on the 5% level of significance. RStudio software (Version 4.3.1; R Core Team, 2023) was used for all statistical analyses, and logistic regression models were fit using the logistf package [22].

### A2) Exploratory phenotyping study

To explore phenotypic differences between FMS mothers with and without autistic children, we utilised an established research study involving detailed phenotyping of 59 patients with moderate or severe FMS who had been identified from the pain management program registry hosted by the same hospital as the service evaluation (Study A1). Study methods and phenotyping results from a subgroup have been published [30]. With few exceptions, these patients were recruited by a senior trainee in pain medicine (RB), and none of these patients’ data had been part of the service evaluation. Recruitment details are summarised in the supplementary Appendix A. The trainee asked participants about the health of their offspring as part of the assessment procedure, and, like the consultant in the service evaluation, did not specifically ask about neurodivergence. If patients volunteered the presence of a neurodivergent condition in offspring, this was documented. Details of the source of the autism diagnosis were not requested. Patients also completed an array of questionnaires and underwent mechanical quantitative sensory testing (Supplementary Materials; Appendix A).

#### Statistical analysis

This sample was split into 3 groups: FMS mothers with autistic children, FMS mothers with non-autistic children, and FMS females without children. To explore differences in pain phenotype between these groups, Kruskal-Wallis tests were conducted, and where a statistically significant difference was found (two-sided 5% level), uncorrected post-hoc comparisons were performed. As this was an exploratory investigation, no correction for testing multiple parameters was applied.

RStudio software (Version 4.3.1; R Core Team, 2023) was used for all statistical analyses.

### B) Online Survey

We conducted an online survey amongst participants of UK pain online support groups aiming to i) record the frequency of autistic children of patients with FMS and other chronic pain conditions in a wider geographical region in the UK, and ii) explore which maternal factors are associated with having an autistic child. We hypothesised that a) there would be an increased rate of autistic children of mothers with FMS compared to mothers with other chronic pain conditions; and b) this relationship would remain even after accounting for maternal or familial history of neurodivergent conditions. The survey was hosted by Qualtrics Survey Software (Qualtrics, 2024) and was advertised first between May 2023 and June 2023, and then between November 2024 and February 2025, through online chronic pain support groups and social media platforms targeting individuals suffering from FMS and general chronic pain. This included *The Pain Relief Foundation* (https://painrelieffoundation.org.uk) and *Pain UK* (https://painuk.org). The advertisement briefly outlined that the study was investigating *neurodivergent* conditions in people with chronic pain and their families, without specifically mentioning autism.

#### Participants

The inclusion criteria were adults with a chronic pain diagnosis who were also parents of children of any age, based in the UK, and fluent in English. Individuals who were pregnant at the time were excluded from participation. Respondents were asked to confirm a chronic pain diagnosis from a specified clinician, although no proof of diagnosis was required. All participants were requested to provide consent and confirm eligibility before starting the online study. Upon completion, participants had the choice to anonymously enter a prize draw to win a £50 Amazon voucher (one for every 50 respondents who entered). The study obtained ethical approval from the University of Liverpool Research Ethics Committee (25^th^ April 2023; Ref: 12195).

#### Questionnaire

The questionnaire was created by the researchers (AH, NF, AG). It recorded mothers’ current pain and maternal/family factors that may be associated with child neurodiversity. Respondents were asked *“Do you have a child with a neurodivergent condition*?”, and if yes, the participant summarised the diagnoses of their neurodiverse child/children in a free text box and provide the number of children affected. For each neurodiverse child, they were asked additional questions regarding each child’s age, whether the child’s other parent had a chronic pain condition at conception/during pregnancy, the mother’s pain before and during pregnancy, and the presence and nature of birth complications. For respondents with more than one neurodiverse child, a summary of the children’s neurodiversity conditions was available through the free text box, however, no data was obtained allowing assignment of these diagnoses to each respective sibling (Appendix B).

#### Data diagnostics

The study was advertised online, therefore, a screening process was undertaken to remove fake responders from the analysis. The exclusion criteria removed: duplicate responses, bots identified by a function of the survey software, nonsensical timelines, non-English responses, and reporting diagnoses which were not related or relevant to the concept (e.g., reporting “headaches” as a neurodivergent condition, and simply “sore” as a chronic pain condition).

Respondents who reported having a child who had *suspected* autism, or was *on a waiting list* for diagnosis, were recorded but not classified as a case of autism. These cases were treated as a control in the analysis.

#### Statistical analysis

Penalised likelihood hierarchical logistic regression analyses were conducted to quantify the association between maternal chronic pain (FMS/Other chronic pain) and autistic children, whilst accounting for the mother’s neurodivergence and the incidence of neurodivergent conditions in the mother’s first-degree family members. A hierarchical regression model was chosen to determine whether the variables of interest improved the model’s ability to predict the binary outcome, after accounting for all other predictor variables. Maternal neurodivergence (diagnosis/no diagnosis) and family neurodivergence (yes/no) were entered into the first model, as inherited and genetic factors are known to play a major role in the aetiology of autism [21]. A second model was created by adding the variable maternal chronic pain condition (FMS/Other chronic pain) to the first model. The association between the individual predictors and the outcome of having a child with an autism diagnosis was determined using ORs and the associated 95% CIs. The model fit was assessed by conducting a model likelihood ratio (LR) chi-square test, and Akaike Information Criterion (AIC) values are also reported.

RStudio software (Version 4.3.1; R Core Team, 2023) was used for all statistical analyses, and logistic regression models were fit using the logistf package [22].

### C) Clinical Prospective Investigation

A second online study invited all patients whose clinic letters had been included in the service evaluation (Study A1) to complete an online questionnaire. Our objectives for this study were to i) expand upon initial data from Study A1 by collecting more comprehensive information on respondents with autistic children, including any maternal contributing factors, and ii) gain further information regarding children’s autistic traits to supplement the parents’ binary report (diagnosis/no diagnosis).

#### Participants

All chronic pain patients included in the clinical service evaluation (N= 589, Study A1), except those who were pregnant at the time of approach, were invited to participate in an online questionnaire. Patients had the choice to enter a prize draw to win a £75 Amazon voucher upon completing the questionnaire. The study received ethical approval from the NHS Health Research Authority Research Ethics Committee Wales REC 2 (Ref: 24/WA/0056).

#### Materials

An online questionnaire was created jointly with an autism specialist (FH) while incorporating Patient and Public Involvement (PPI) in its development. Chronic pain patients involved in an existing FMS PPI group at our research institution advised on the content of the survey, its clarity, and sensitivity. Several validated scales were included in the questionnaire. Internal reliability was assessed using McDonald’s ω, which was calculated from the current study sample. Data were collected using Qualtrics Survey Software (Qualtrics, 2024). See the supplementary materials for the full questionnaire (Appendix C).

1. The General Anxiety Disorder (GAD-7; [42]) is a 7-item *self-report* scale which was developed to measure general anxiety disorder. This scale was included based on the consideration that higher maternal anxiety may relate to an increased desire to have clarity about their child’s diagnosis, potentially increasing the probability of obtaining an autism diagnosis. Responses to all items are scored on a 4-point Likert scale ranging from *0 (not at all)* to *3 (nearly every day)* in reference to the previous 2 weeks. Total scores range from 0 to 21, with higher scores indicating higher severity of general anxiety. A score of ≥10 is considered the cut-off for moderate anxiety. The overall scale had excellent internal reliability (McDonald’s ω_t_ = 0.97).
2. The Autism Spectrum Quotient Adult (AQ-10 Adult; [1]) is a 10-item *self-report* scale developed as a brief screening tool to capture autistic traits in adults. Total scores range from 0 to 10. A cut-off value of ≥6 indicates a possible case of autism. The overall scale had good internal reliability in the present sample (McDonald’s ω_t_ = 0.80).
3. The Ritvo Autism Asperger Diagnostic Scale (RAADS-14; [16]) is a 14-item *self-report* scale which is designed to assess traits associated with autism in adults. The scale consists of 3 subscales including mentalising deficits, social anxiety, and sensory reactivity. All items, except item 6, which is reversed, are scored *3 (true now and when I was young), 2 (true only now), 1 (true only when I was young),* and *0 (never true).* Total scores range from 0 to 42 with a cut-off value of ≥14 being indicative of possible autism. The overall scale had acceptable internal reliability (McDonald’s ω_h_ = 0.76).
4. Parent-Report Autism Spectrum Quotient (Parent-report AQ-10; [1]) is a 10-item scale developed for parents to complete *referring to their child*. Total scores range from 0 to 10, and a cut-off value of ≥6 is indicative of possible autism. The overall scale had good internal reliability (McDonald’s ω_t_ = 0.87).
5. The Social Communication Questionnaire – Lifetime (SCQ; [41]) is a 40-item, parent-report measure for screening symptomology associated with autism. All items are answered dichotomously *(yes/no)*. Item 1 records whether the referred child has the ability to speak in short phrases or sentences, and items 2 – 40 are scored and summed. Total scores range from 0 to 39 with a cut-off value of ≥15 suggesting autism. The overall scale had excellent internal reliability (McDonald’s ω_t_ = 0.91).

#### Procedure

After providing consent and passing eligibility checks, patients were asked about their chronic pain history and general health, and to complete 3 standardised questionnaires referring to themselves (GAD-7, AQ-10 Adult, RAADS-14). After this, patients were asked to complete questions referring to the health of all their children, in birth order. This included the parent-report AQ10 *for each of their children*. The SCQ was then only completed for children who respondents either reported as having an autism diagnosis, or about whom they had concerns regarding their social communication, behaviour, and/or development. See supplementary materials for details about children’s AQ10 and SCQ scores (Supplementary Table 14 and 15; Appendix C).

Patients with autistic children were asked further questions regarding any complications during pregnancy, delivery or after birth, co-occurring child health conditions, child intellectual disability, and the level of any child support needs assigned by a doctor in reference.

#### Statistical analysis

Hierarchical penalised likelihood logistic regression analyses were conducted to test the association of maternal chronic pain condition (FMS/ Other chronic pain) whilst accounting for family history variables (autistic mothers and the incidence of neurodivergent conditions in first-degree family members) and psychological variables (GAD-7 scores). Maternal autism diagnosis status (diagnosis/ no diagnosis) and family neurodivergence (yes/no) were entered into the first model, GAD-7 scores were added into the second model, and maternal chronic pain condition was entered into the third model. The association between these individual parameters and the outcome of having a child with an autism diagnosis was determined using ORs with 95% CIs. The model fit was determined by conducting a model LR chi-square test and AIC values were reported. RStudio software (Version 4.3.1; R Core Team, 2023) was used for all statistical analyses. Logistic regression models were fit using the logistf package [22], and internal reliability was assessed using the psych package [39].

## Results

### A1) Clinical Service Evaluation

The cohort included 589 total patients, 398 (67.57%) females and 191 (32.43%) males. The consultant had documented an enquiry about the health of the respective patient’s offspring in 89% of cases. Among the patients with confirmed biological children, 31 (8.68%) patients had at least one child with an autism diagnosis. Thirteen (16.25%) mothers with FMS, 4 mothers with CRPS (14.81%) and 6 (4.05%) mothers with other chronic pain conditions reported having at least one autistic child. Table 1 displays the full sample by chronic pain condition.

**Table 1.** The total sample of chronic pain patients by condition and the frequency of reporting children with an autism diagnosis.

| **Chronic pain condition** | **Total N of patients (females, %)** | **Patients with confirmed biological children (females, %)** | **Patients with at least one autistic child N[%]<sup>a</sup>** | **Female patients with at least one autistic child N[%]<sup>b</sup>** | **Male patients with at least one autistic child N [%]<sup>c</sup>** |
| --- | --- | --- | --- | --- | --- |
| *FMS* | 119 (107 females, 89.92%<sup>d</sup>) | 86 (80, 93.02%<sup>d</sup>) | 13 [15.12%] | 13 [16.25%] | 0 [0.00%] |
| *CRPS* | 67 (42, 62.69%) | 38 (26, 68.42%) | 7 [18.42%] | 4 [14.81%] | 3 [25.00%] |
| *Other chronic pain conditions* | 403 (249, 61.79%) | 239 (148, 61.92%) | 11 [4.60%] | 6 [4.05%] | 5 [5.56%] |

<sup>a</sup> Patients with at least one autistic child as a percentage of those with confirmed biological children within the chronic pain condition category.
<sup>b</sup> Female patient with at least one autistic child as a percentage of females with confirmed biological children within the chronic pain condition category.
<sup>c</sup> Male patient with at least one autistic child as a percentage of males with confirmed biological children within the chronic pain condition category.
<sup>d</sup> 3 of these female patients had both FMS and CRPS (not listed in the CRPS group), 1 of whom had an autistic child.

Information about the onset of mothers’ pain in relation to their children’s ages/ birth was not consistently documented; of the available data, 3 mothers with FMS (30.00%) and 2 mothers with CRPS (100.00%) reported that the onset of their FMS or CRPS symptoms, respectively, occurred before childbirth. Two mothers with other chronic pain (33.3%) reported that they had pain before childbirth (Table 2). Table 2 describes further available information pertaining to children with an autism diagnosis.

**Table 2.**
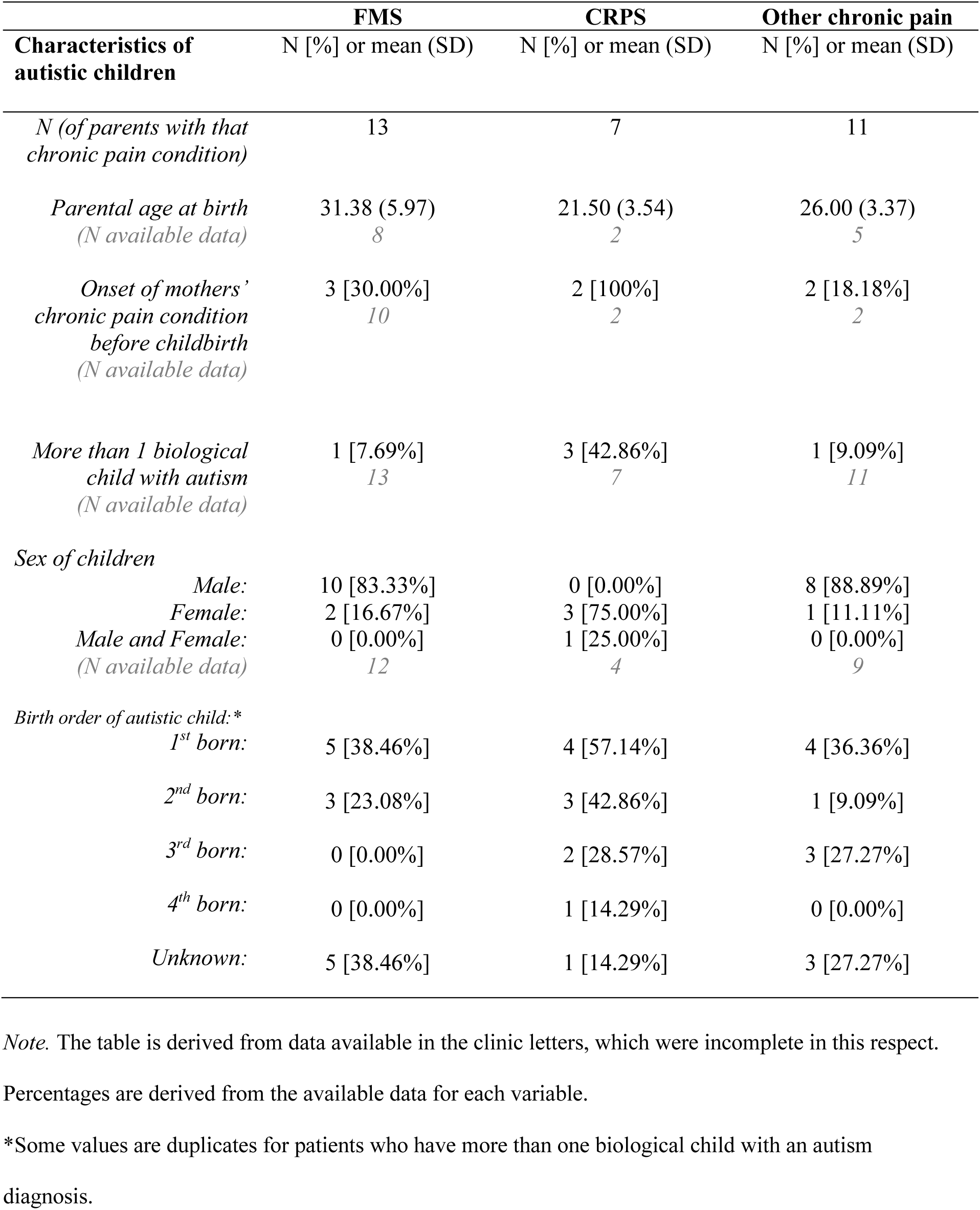
Characteristics of autistic children, by parental chronic pain condition in the clinical service evaluation.

| **Characteristics of autistic children** | **FMS**<br>N [%] or mean (SD) | **CRPS**<br>N [%] or mean (SD) | **Other chronic pain**<br>N [%] or mean (SD) |
| --- | --- | --- | --- |
| *N (of parents with that chronic pain condition)* | 13 | 7 | 11 |
| *Parental age at birth*<br>*(N available data)* | 31.38 (5.97)<br>8 | 21.50 (3.54)<br>2 | 26.00 (3.37)<br>5 |
| *Onset of mothers' chronic pain condition before childbirth*<br>*(N available data)* | 3 [30.00%]<br>10 | 2 [100%]<br>2 | 2 [18.18%]<br>2 |
| *More than 1 biological child with autism*<br>*(N available data)* | 1 [7.69%]<br>13 | 3 [42.86%]<br>7 | 1 [9.09%]<br>11 |
| *Sex of children* |  |  |  |
| *Male:* | 10 [83.33%] | 0 [0.00%] | 8 [88.89%] |
| *Female:* | 2 [16.67%] | 3 [75.00%] | 1 [11.11%] |
| *Male and Female:*<br>*(N available data)* | 0 [0.00%]<br>12 | 1 [25.00%]<br>4 | 0 [0.00%]<br>9 |
| *Birth order of autistic child: \** |  |  |  |
| *1<sup>st</sup> born:* | 5 [38.46%] | 4 [57.14%] | 4 [36.36%] |
| *2<sup>nd</sup> born:* | 3 [23.08%] | 3 [42.86%] | 1 [9.09%] |
| *3<sup>rd</sup> born:* | 0 [0.00%] | 2 [28.57%] | 3 [27.27%] |
| *4<sup>th</sup> born:* | 0 [0.00%] | 1 [14.29%] | 0 [0.00%] |
| *Unknown:* | 5 [38.46%] | 1 [14.29%] | 3 [27.27%] |

*Note.* The table is derived from data available in the clinic letters, which were incomplete in this respect.
Percentages are derived from the available data for each variable.
\*Some values are duplicates for patients who have more than one biological child with an autism diagnosis.

Although not classified as a case of autism in the analysis, 7 patients reported that they had one child who they *suspected* was autistic, but no diagnosis (2 FMS; 1 CRPS; 4 other chronic pain).

Figure 1a shows the results of the penalised logistic regression model examining the association between maternal chronic pain condition and the likelihood of children with autism diagnoses. The overall model was statistically significant χ^2^(2) = 11.00, *p*=.004, McFadden R^2^ =0.01. The likelihood of autistic offspring was estimated at OR: 4.38 (95% CI: 1.70 –12.45) for maternal FMS and at OR: 4.38 (1.14 - 15.70) for maternal CRPS, compared to other chronic pain.

**Figure 1.**
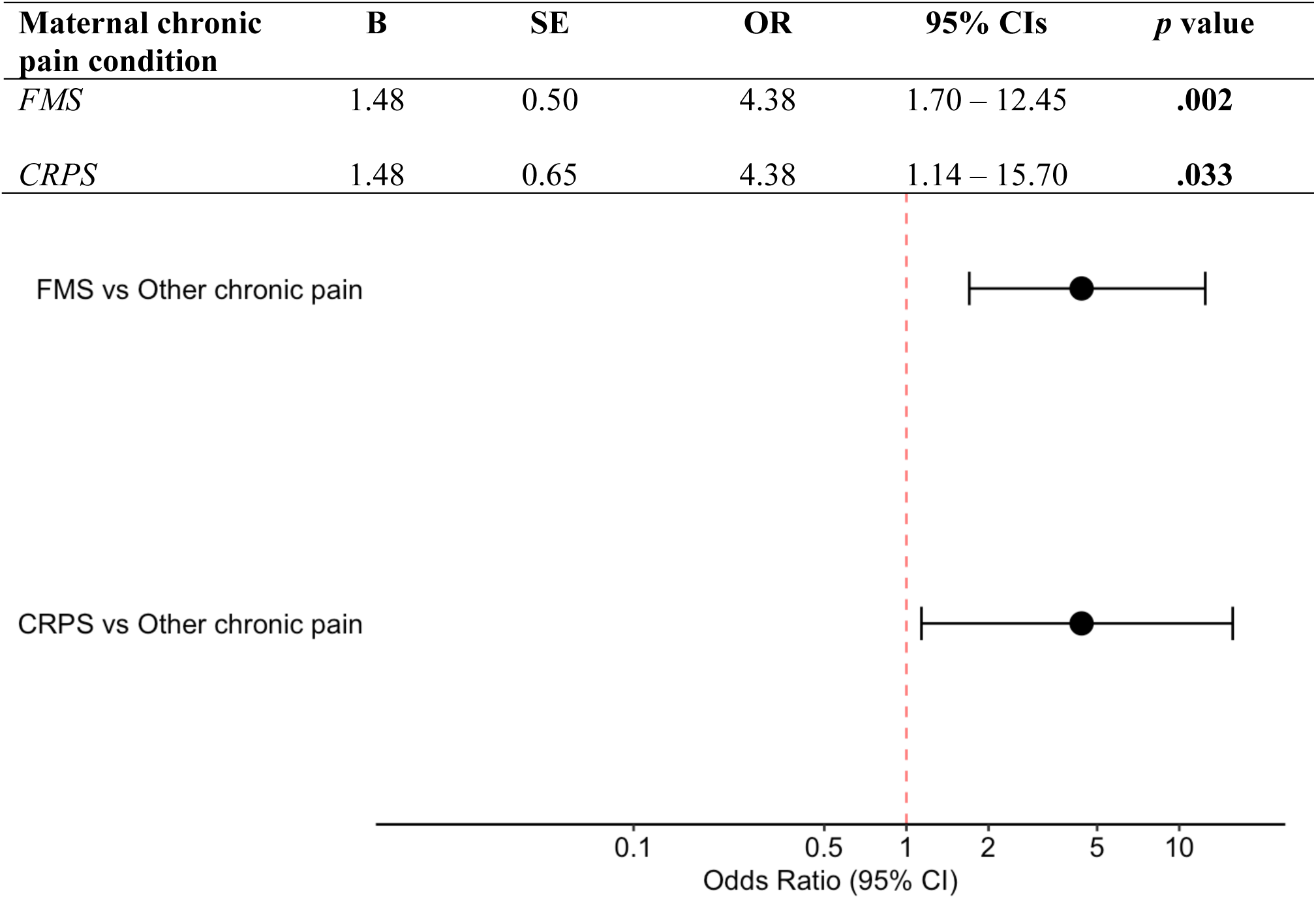
(a) Penalised logistic regression analysis showing the association of maternal FMS and CRPS, relative to other chronic pain conditions, to children with an autism diagnosis. (b) Forest plot showing ORs and 95% CIs.

### A2) Exploratory phenotyping study

Five/44 (11.36%) FMS participant mothers reported having an autistic child. In comparison to the mothers with non-autistic children, mothers of autistic children had statistically significantly lower scores on the Widespread Pain Index (i.e., their pain was less widespread, median 10.00, IQR 2.00 vs. median 15.00, IQR 5.00; *p* =.027, η^2^ = 0.09 uncorrected Kruskal-Wallis test) and on the Short-Form McGill Pain Questionnaire *intermittent* subscale (i.e., lower reported severity of intermittent pain, median 3.40, IQR 2.02 vs. median 5.80, IQR 4.25; *p* =.049, η^2^ = 0.07). There were no other significant differences between these 2 groups on any of the measured demographic, impact, pain, or sensory parameters (Supplementary Table 2; Appendix A).

### B) Online Survey

This cohort of parents included 710 participants (589 FMS, 121 other chronic pain), of whom 622 were mothers (87.61%) and 80 were fathers (11.27%). Eight respondents (one of whom reported having a child with an autism diagnosis) did not provide their sex and, therefore, were not included in the analysis. One hundred and thirty-seven (25.95%) mothers with FMS and 10 (10.64%) mothers with other chronic pain self-reported at least one child with an autism diagnosis.

In the “other chronic pain” cohort, the most common pain conditions were back pain, arthritis and neck/shoulder pain; two respondents reported having pain due to an autoimmune condition (Supplementary Table 3; Appendix B).

Table 3 describes the participant characteristics of the mothers-only sample, stratified by maternal chronic pain condition and the autism diagnosis status of children. Although not classified as a case of autism in the analysis, 7 mothers with FMS reported having a child they *suspected* was autistic or indicated that they were on a waiting list for diagnosis.

**Table 3.**
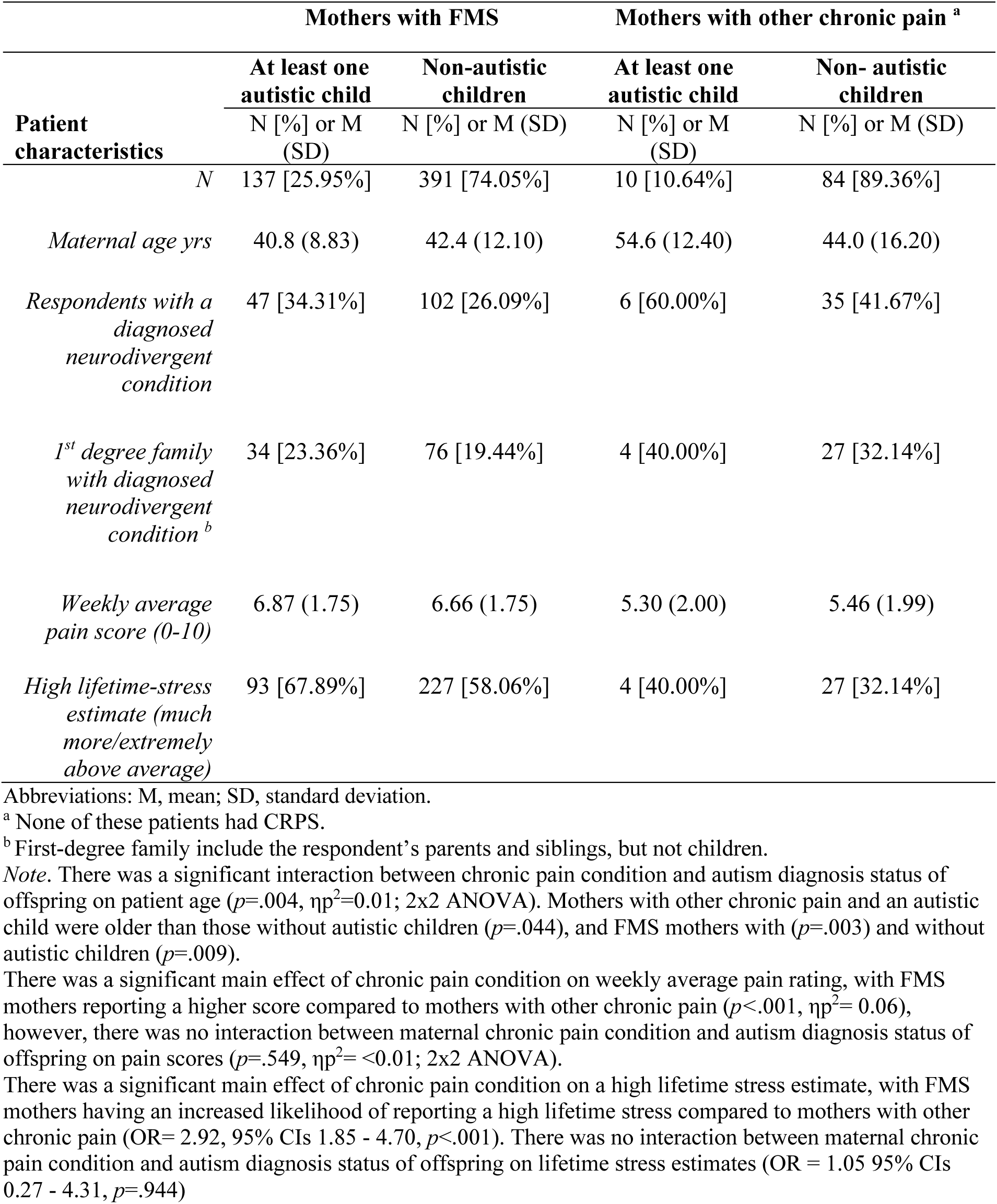
Characteristics of mothers with FMS or other chronic pain, stratified by the autism diagnosis status of children.

| Patient characteristics | Mothers with FMS |  | Mothers with other chronic pain <sup>a</sup> |  |
| --- | --- | --- | --- | --- |
|  | At least one autistic child | Non-autistic children | At least one autistic child | Non- autistic children |
|  | N [%] or M (SD) | N [%] or M (SD) | N [%] or M (SD) | N [%] or M (SD) |
| *N* | 137 [25.95%] | 391 [74.05%] | 10 [10.64%] | 84 [89.36%] |
| *Maternal age yrs* | 40.8 (8.83) | 42.4 (12.10) | 54.6 (12.40) | 44.0 (16.20) |
| *Respondents with a diagnosed neurodivergent condition* | 47 [34.31%] | 102 [26.09%] | 6 [60.00%] | 35 [41.67%] |
| *1<sup>st</sup> degree family with diagnosed neurodivergent condition <sup>b</sup>* | 34 [23.36%] | 76 [19.44%] | 4 [40.00%] | 27 [32.14%] |
| *Weekly average pain score (0-10)* | 6.87 (1.75) | 6.66 (1.75) | 5.30 (2.00) | 5.46 (1.99) |
| *High lifetime-stress estimate (much more/extremely above average)* | 93 [67.89%] | 227 [58.06%] | 4 [40.00%] | 27 [32.14%] |

Abbreviations: M, mean; SD, standard deviation.
<sup>a</sup> None of these patients had CRPS.
<sup>b</sup> First-degree family include the respondent's parents and siblings, but not children.
*Note.* There was a significant interaction between chronic pain condition and autism diagnosis status of offspring on patient age ( $p=.004$ , $\eta^2=0.01$ ; 2x2 ANOVA). Mothers with other chronic pain and an autistic child were older than those without autistic children ( $p=.044$ ), and FMS mothers with ( $p=.003$ ) and without autistic children ( $p=.009$ ).
There was a significant main effect of chronic pain condition on weekly average pain rating, with FMS mothers reporting a higher score compared to mothers with other chronic pain ( $p<.001$ , $\eta^2= 0.06$ ), however, there was no interaction between maternal chronic pain condition and autism diagnosis status of offspring on pain scores ( $p=.549$ , $\eta^2= <0.01$ ; 2x2 ANOVA).
There was a significant main effect of chronic pain condition on a high lifetime stress estimate, with FMS mothers having an increased likelihood of reporting a high lifetime stress compared to mothers with other chronic pain (OR= 2.92, 95% CIs 1.85 - 4.70, $p<.001$ ). There was no interaction between maternal chronic pain condition and autism diagnosis status of offspring on lifetime stress estimates (OR = 1.05 95% CIs 0.27 - 4.31, $p=.944$ )

Three (5.26%) fathers with FMS and 1 (4.35%) father with other chronic pains reported having an autistic child. Additional analyses showed a statistically significant main effect of parental sex on the likelihood of autism diagnoses in children. The likelihood of autistic offspring was estimated at OR 4.82 (95% CI: 2.03 – 14.85, *p* <.001) for mothers compared to fathers. There was no significant interaction between parental sex and chronic pain condition (*p* =.341). Supplementary Table 5 shows further details about the fathers-only sample (Appendix B).

Patients also reported other neurodivergent conditions in their children. Overall, ADHD was the second most reported at a rate of 21.78% for FMS mothers and 14.89% for mothers with other chronic pain (non-significant difference, supplementary Table 6; Appendix B). Different neurodivergent conditions were rarely noted (Supplementary Table 4; Appendix B).

Table 4 shows, for mothers with only one child with autism, their age at birth of the autistic child, the proportion with a pain diagnosis, or pain symptoms consistent with symptoms at survey completion, and their birth complications with the autistic child. Note that this information was only available for mothers with one neurodiverse child (N=101/137).

**Table 4.** Characteristics of the mothers and their autistic child, stratified by maternal chronic pain condition.

| **Characteristics of mother and autistic child** | **FMS** | **Other chronic pain** |
| --- | --- | --- |
|  | N [%] or mean (SD) | N [%] or mean (SD) |
| *N of mothers with one autistic child* | 101 | 7 |
| *Maternal age at birth* | 26.69 (5.26) | 28.00 (5.02) |
| *Maternal (current) chronic pain <u>diagnosis</u> before/around childbirth* | 25 [24.75%] | 1 [14.29%] |
| *Onset of maternal (current) chronic pain <u>symptoms</u> before/around childbirth* | 43 [42.57%] | 3 [42.86%] |
| *If (maternal) pain during pregnancy: average pain intensity (0-10) <sup>a</sup>* | 6.14 (1.99) | 4.00 (3.27) |
| *If (maternal) pain before pregnancy: average pain intensity (0-10) <sup>a</sup>* | 5.16 (1.77) | 3.00 (3.16) |
| *Child's father had a known chronic pain condition around conception/pregnancy* | 22 [21.78%] | 1 [14.29%] |
| *Child with autism co-occurring with other neurodivergent conditions* | 7 [6.93%] | 2 [28.57%] |
| *Birth/ pregnancy complications* | 51 [50.50%] | 1 [14.29%] |
| - *Gestational age <35 wks* | 11 [10.89%] | 0 [0.00%] |
| - *Low birth weight* | 14 [13.86%] | 0 [0.00%] |
| - *Birth injury/trauma* | 17 [16.83%] | 0 [0.00%] |
| - *Birth defects associated with the CNS* | 4 [3.96%] | 0 [0.00%] |
| - *Meconium aspiration syndrome* | 2 [1.98%] | 1 [14.29%] |
| - *Neonatal or epileptic encephalopathy* | 1 [0.99%] | 0 [0.00%] |
| - *Genetic disorder* | 2 [1.98%] | 0 [0.00%] |

*Note.* “Current” = respondent’s reported chronic pain conditions (FMS or other chronic pain) at the time of the survey. The presence of the maternal pain diagnosis or of corresponding symptoms around the time of pregnancy were calculated from the child’s age and the respondent’s reported year of diagnosis and of (yet undiagnosed) pain onset.
<sup>a</sup> For mothers with FMS, *pain before/during pregnancy* refers to the presence of widespread pain; for mothers with other chronic pain, this refers to the presence of any pain.

Figure 2a shows the results of the penalised hierarchical logistic regression model examining the association between maternal variables and the likelihood of children with autism diagnoses. The first model, including the family-history factors, maternal neurodivergence and first-degree family neurodivergence, was non-significant χ²(2) = 2.93, *p* =.231, AIC = 672.78, McFadden’s R^2^= 0.01.

**Figure 2.**
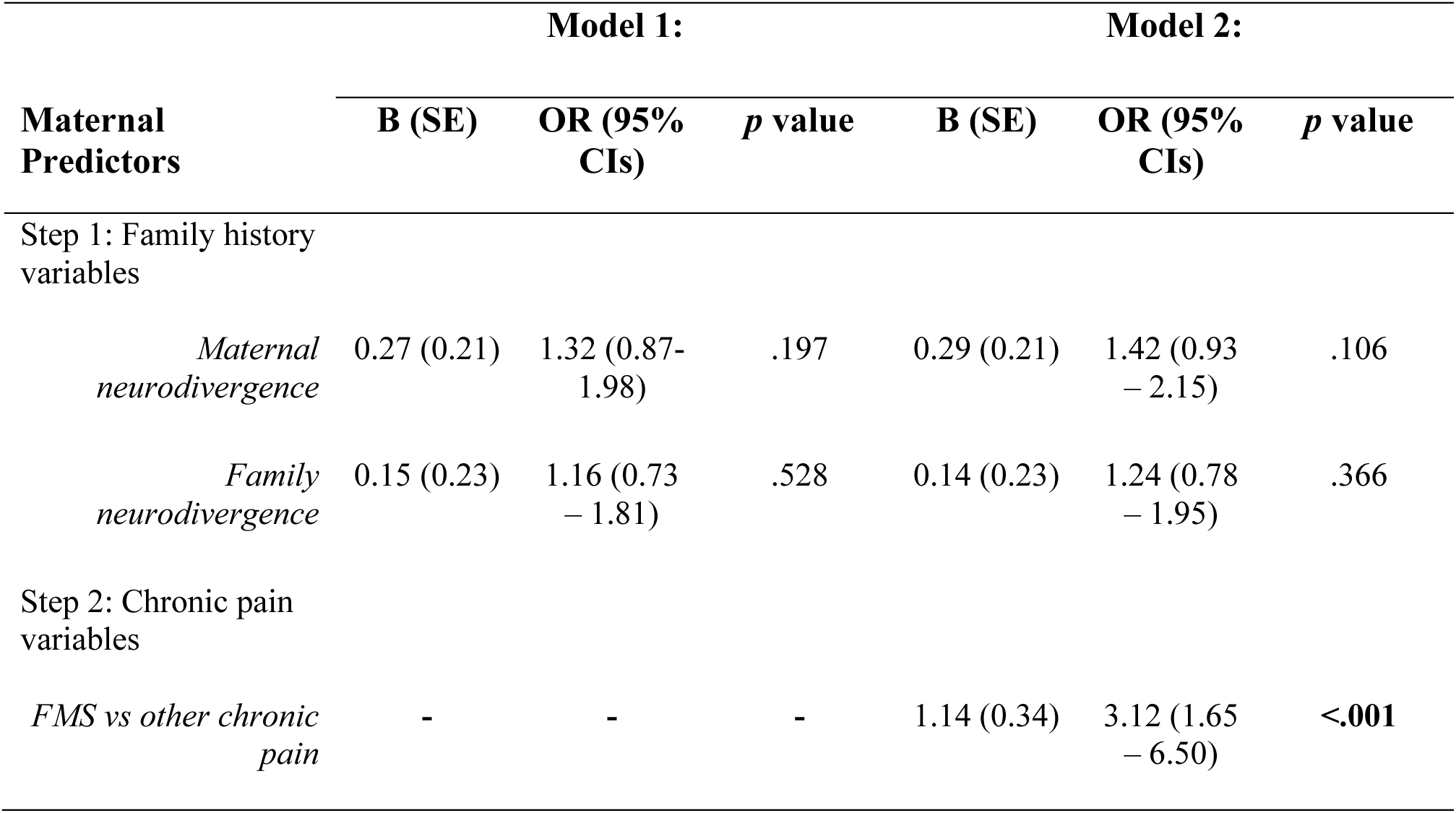
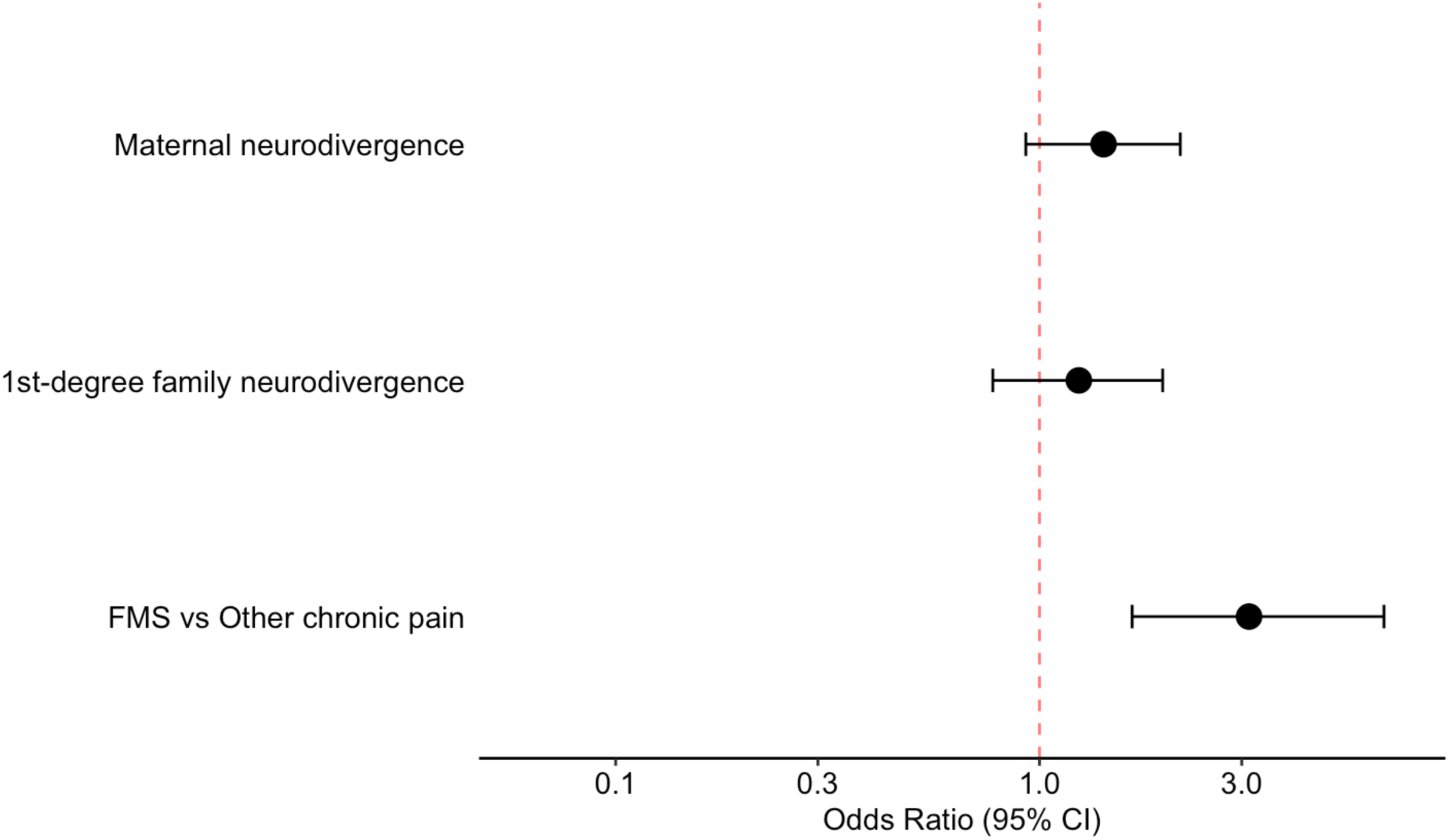
(a) Penalised logistic regression analysis showing the association of maternal variables to children with an autism diagnosis. (b) Forest plot showing ORs and 95% CIs.

Adding maternal chronic pain condition into the second model significantly improved the model fit relative to the family-history only model χ²(1)= 13.57, *p*<.001. With this, the overall model was statistically significant χ²(3) = 16.50, *p*<.001, AIC = 672.12, McFadden’s R^2^= 0.02. The likelihood of autistic offspring was estimated at OR 3.12 (95% CI: 1.65 – 6.50) for mothers with FMS compared to mothers with other chronic pain.

### C) Clinical Prospective Investigation

The cohort included a total of 66 patients (50 [75.76%] female, 16 [24.24%] male); the overall response rate was 11.2% (24.6% FMS, 17.7% CRPS, 6.3% other chronic pain). Nineteen of the 24 mothers with FMS (79.16%) had an FMS diagnosis during the service evaluation, all 7 mothers with CRPS (100%), and 8 out of the 11 mothers with other chronic pain (72.73%) were categorised as other chronic pain in the service evaluation.

Among the maternal sample, 8 mothers with FMS reported having an autistic child (33.33%), whereas no cases were reported among mothers with other chronic pain. Table 5 shows the characteristics of the maternal sample, stratified by the autism diagnosis status of offspring, and Table 6 shows the characteristics of FMS mothers and their autistic child. Responding mothers with CRPS were not included in the analysis due to a small sample size (N= 7), see supplementary materials for descriptive details about CRPS mothers (Supplementary Table 10; Appendix C).

**Table 5.** Characteristics of mothers with FMS and other chronic pain, stratified by the autism diagnosis status of children.

| Patient characteristics | Mothers with FMS |  | Mothers with other chronic pain |
| --- | --- | --- | --- |
|  | Autistic child | Non-autistic child | Non-autistic child |
|  | N [%] or median (IQR) | N [%] or median (IQR) | N [%] or median (IQR) |
| *N* | 8 [33.33%] | 16 [66.67%] | 11 [100%] |
| *Maternal age yrs (mean (SD))* | 48.8 (7.34) | 49.3 (8.93) | 49 (14.2) |
| *Autistic respondent* | 0 [0.00%] | 2 [12.5%] | 0 [0.00%] |
| *1<sup>st</sup> degree family with diagnosed neurodivergent condition <sup>a</sup>* | 3 [37.50%] | 1 [6.25%] | 2 [18.18%] |
| *GAD-7 (0-21)* | 7 (8) | 7.5 (11.2) | 13 (8.5) |
| *Scored above cutoff ($\geq 10$)* | 3 [37.50%] | 7 [43.75%] | 7 [63.64%] |
| *RAADS-14 Total (0-42)* | 25 (14.5) | 20.5 (20) | 24 (16) |
| *Scored above cutoff ($\geq 14$)* | 6 [75.00%] | 10 [62.50%] | 9 [81.82%] |
| *AQ-10 (0-10)* | 4.5 (1.25) | 4.5 (1.25) | 5 (1.5) |
| *Scored above cutoff ($\geq 6$)* | 2 [25.00%] | 4 [25.00%] | 3 [27.27%] |

Abbreviations: IQR, Interquartile Range; SD, standard deviation.
<sup>a</sup>First-degree family include the respondent's parents and siblings, but not children.
*Note.* There were no significant differences in age ( $p=.969$ $\eta^2= <0.01$ ; ANOVA), AQ-10 scores ( $p=.976$ , $\eta^2= -0.06$ ), or RAADS-14 scores ( $p=.757$ , $\eta^2= -0.05$ ; Kruskal-Wallis) between groups.

**Table 6.** Characteristics of autistic children, of mothers with FMS.

| Characteristics of autistic child |  | Mothers with FMS<br>N [%] or mean (SD) |
| --- | --- | --- |
| *N (of mothers with FMS)* |  | 8 |
| *Maternal age at birth*<br>*(N available data)* |  | 25.90 (6.62)<br>8 |
| *AQ-10 score median (IQR)(0-10; cutoff ≥6)*<br>*(N available data)* |  | 8.50 (1.50)<br>8 |
| *SCQ score median (IQR) (0-39; cut off ≥15)*<br>*(N available data)* |  | 24.00 (6.50)<br>7 |
| *Age at diagnosis yrs*<br>*(N available data)* |  | 14.29 (4.82)<br>7 |
| *Sex = male*<br>*(N available data)* |  | 4 [50.00%]<br>8 |
| *Birth order:* | *1<sup>st</sup> child:* | 3 [37.50%] |
|  | *2<sup>nd</sup> child:* | 2 [25.00%] |
|  | *3<sup>rd</sup> child:* | 3 [37.50%] |
|  | *(N available data)* | 8 |
| *Any maternal chronic pain during pregnancy*<br>*(N available data)* |  | 4 [50.00%]<br>8 |
| *Maternal FMS or widespread pain during pregnancy*<br>*(N available data)* |  | 2 [28.57%]<br>7 |
| *Birth/ pregnancy complications*<br>*(N available data)* |  | 2 [28.57%]<br>7 |
|  |  | • Respiratory distress syndrome<br>• Low blood sugar |
| *Co-occurring intellectual disability*<br>*(N available data)* |  | 3 [42.86%]<br>7 |
| *Level of support needs:* |  |  |
| *Level 1 (requiring support):* |  | 0 [0.00%] |
| *Level 2 (requiring substantial support):* |  | 3 [100.00%] |
| *Level 3 (requiring very substantial support):*<br>*(N available data)* |  | 0 [0.00%]<br>3 |
| *Child's other co-occurring conditions* |  | • Pulmonary stenosis<br>• Cerebral Palsy<br>• Asthma (x2)<br>• ADHD |

*Note.* Parents did not know or report answers for all the presented variables. Percentages are derived from the available data for each variable.

In all cases, patients reported having only one autistic child. All autistic children included in the analysis scored above the recommended cut-off values on the parent-report AQ10 (score of ≥6) and SCQ (score of ≥15). See Table 6 for further information pertaining to autistic children.

Figure 3a shows the results of the hierarchical penalised likelihood logistic regression model examining the association between maternal variables and the offspring with autism diagnoses. The first model, including family history variables: maternal autism diagnosis and first-degree family neurodivergence, was not statistically significant χ^2^(2) = 3.53, *p*=.171, AIC = 40.99, McFadden’s R^2^ =-0.03.

**Figure 3.**
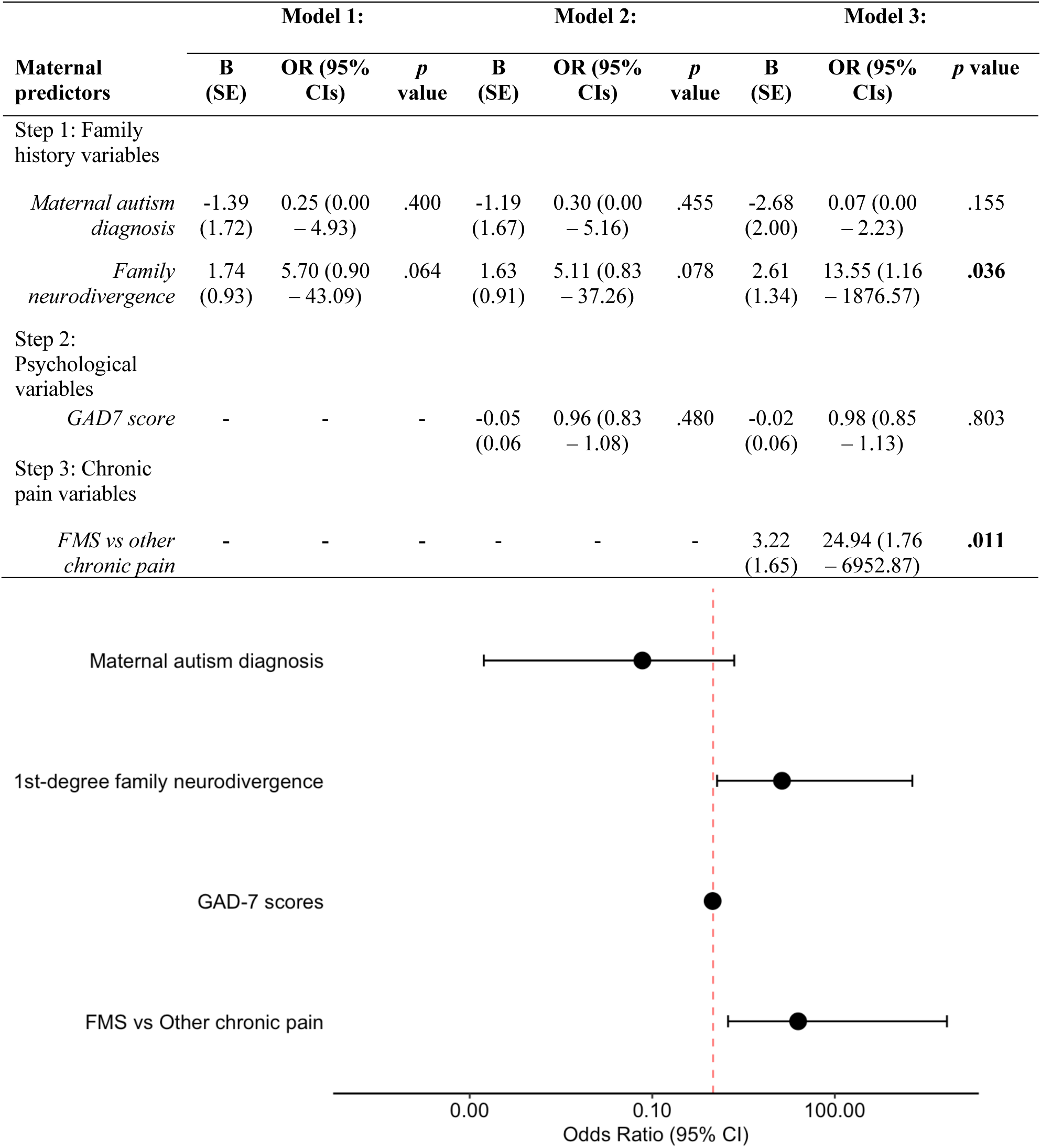
(b) Penalised logistic regression analysis showing the association of maternal variables to children with an autism diagnosis. (b) Forest plot showing ORs and 95% CIs.

The model LR chi-square test indicated that adding the psychological variable, GAD-7 scores, did not significantly improve the model fit χ^2^(1) = 0.54 *p*=.539, and this second model was not statistically significant χ^2^(3) = 3.91, *p*=.271, AIC = 39.11, McFadden’s R^2^ = 0.12.

The third model included maternal chronic pain condition (FMS and other chronic pain). The model LR chi-square test suggested that adding these variables significantly improved the model fit χ^2^(1) = 6.27, *p*=.012. With this, the overall model was statistically significant χ^2^(4) = 10.18, *p*=.038, AIC = 42.60, McFadden’s R^2^ = 0.13. The likelihood of autistic offspring was estimated at OR 24.94 (95% CI: 1.76 – 6952.87) for mothers with FMS compared to mothers with other chronic pain. The likelihood for mothers with first-degree neurodivergent relatives was estimated at OR 13.55 (1.16 – 1876.57). However, the precision of these estimates is poor due to a small sample size.

## Discussion

To our knowledge, no previous report has explored the occurrence of autism specifically in the children of patients suffering from FMS. We found an increased likelihood of autistic children with mothers who have FMS compared to mothers with other chronic pain conditions in each of four cohorts.

A clinical service evaluation (Study A1) included all chronic pain patients seen by a consultant at a tertiary centre over 2 years. Mothers with FMS and CRPS reported a high prevalence of autistic children (16% and 15%, respectively) versus patients presenting with other pain conditions (4%). This relationship was found to be significant, with 4.38 increased odds in mothers with FMS, and 4.38 increased odds for mothers with CRPS.

An exploratory phenotyping study (Study A2) analysed the responses of FMS patients who had also been routinely asked about their children’s health. Eleven percent of mothers with FMS reported an autistic child. Together, these findings suggest a much higher prevalence of autistic children of FMS mothers compared to the UK general population estimate of 1%-3% [8; 33]. Comparison of demographic and FMS characteristics between those with and without autistic children in Study A2 revealed a lower reported widespread pain score and less intermittent pain (e.g., shooting, stabbing pain) for mothers with an autistic child. These results should be considered exploratory, however, the moderate effect sizes raise the possibility that there might be a difference in the FMS phenotype in mothers who have an autistic child.

An online survey (Study B) was distributed among members of online pain support groups to assess the frequency of autistic children in FMS mothers, beyond both tertiary healthcare settings and the geographical restriction of the Study A1/2 (NW-England). The aim was to confirm the autism incidence findings and explore mothers’ personal and first-degree family members’ self-reported neurodiversity, as genetic factors play a major role in the aetiology of autism [21]. Consistent with the service evaluation, a higher incidence of autism diagnoses in offspring was reported for mothers with FMS compared to mothers with other chronic pain conditions (26% and 11%, respectively). Astonishingly, although not the primary objective of this investigation we found that 58% of all FMS mothers had at least one child with at least one neurodiversity condition. FMS in mothers remained statistically significantly associated with 3.12 increased odds of autistic children compared to mothers with other chronic pain after accounting for maternal and first-degree familial history of neurodivergence. FMS fathers reported autistic children much less frequently than FMS mothers, and at a similar prevalence across conditions (4-5%). Unexpectedly, mothers with other chronic pain reported diagnosed neurodivergent conditions in themselves and their first-degree family members more frequently than FMS mothers.

The high reported neurodivergence prevalences in responders and their relatives across groups are likely affected by a selection bias whereby those with a known occurrence of neurodivergence in themselves or their family are more likely to participate. This high prevalence is consistent with current literature suggesting an overlap between neurodivergence and chronic pain [9; 29], however, the increased prevalence in FMS mothers compared to others suggests previously-unreported variability within the chronic pain group. Biological fathers might provide a greater genetic contribution in this group – we did not assess the fathers’ or paternal first-degree family members’ neurodiversity in this study, but this may be unlikely given the low offspring autism rates reported by participating fathers in the same study. Alternatively, this finding might suggest a lower genetic propensity for the autistic children of FMS mothers than of mothers with other chronic pain conditions. Correspondingly, non-genetic factors, including perinatal factors such as pathogenic autoantibodies [6; 13; 25], or post-natal factors, may be more relevant in these children of FMS mothers. Additional studies will be required to understand this area.

In this online survey, 51% of FMS mothers who had only one child with autism reported having birth/pregnancy complications with their autistic child, with birth injury or trauma (17%), low birth weight (14%), and premature birth (11%) being most reported. This can be aligned with previous research which demonstrates that such parameters are associated with autistic children [4; 28]. Moreover, FMS in pregnancy has been found to be associated with several adverse maternal and neonatal outcomes, including premature birth [31]. In our study, data on birth and pregnancy outcomes were not collected for mothers without neurodivergent children, therefore, it is unclear at present whether the adverse outcomes are specific to FMS mothers with autistic children.

A final clinical prospective study (Study C) invited all patients from the initial service evaluation to complete a survey capturing detailed data on maternal factors and children’s autistic traits. Mothers with FMS, and generally a first-degree family history of neurodivergence were statistically significantly associated with an increased likelihood of autistic children, the latter aligning with the well-documented role of genetics in the development of autism [21]. Previous evidence suggests that FMS patients display high health anxiety [44], which in turn could lead to a heightened awareness regarding their children’s health. The observed lack of association of maternal GAD-7 scores with autism diagnoses in children does not support this factor’s relevance. No significant differences were found across conditions for maternal AQ-10 and RAADS-14 autistic trait measure scores, providing again no signal for a *specific* genetic load in FMS mothers. Notably, regardless of chronic pain category, a substantial proportion of this cohort (67% FMS mothers; 82% other pain mothers) scored above the cut-off for suggested autism in the RAADS-14, but not the AQ10 (25% FMS mothers; 27% other chronic pain mothers). It is possible that certain items on the RAADS-14 such as the second item ‘*some ordinary textures that do not bother others feel very offensive when they touch my skin’* may capture characteristics of chronic pain conditions. Further research will be required to understand this discrepancy.

This is the first study to demonstrate a statistically significant association between maternal FMS and autism specifically in children, when compared to other chronic pain conditions. Our results are consistent with those from Kelly [26] who reported an increased prevalence of neurodivergent conditions among the *relatives* (first-and second-degree) of 13 patients with *FMS and concomitant hypermobility*, as compared to a control group of osteoarthritis patients. Whether the mechanisms underpinning increased autism rates in children of patients with FMS are identical to those explaining increased autism rates in relatives overall is uncertain. The findings of our online survey hint at the possibility that the development of autism in children of FMS mothers might be explained, at least in part, by the perinatal environment rather than solely by genetic factors.

There were several limitations to the current study. First, self-selection bias may have influenced the high reported prevalence of diagnosed autism in Study B and C. However, this is unlikely to account for the disparity between chronic pain conditions. Second, the research relied on parental reports of autism diagnoses. Although parental confirmation of a diagnosis from a clinician was required in the prospective study, this could not be formally verified. All autistic children in the analysis exceeded the recommended cut-off values on the parent-reported AQ10 and SCQ autistic trait measures providing some reassurance about the validity of these parent reports. Third, the online survey relied on self-report of chronic pain, and diagnoses could not be independently verified. Fourth, the online survey did not allow collection of birth/pregnancy information where mothers had more than one child with neurodiversity. Fifth, we did not ask in the online survey whether FMS mothers had any pain in the perinatal period other than that which they had at the survey completion time. Sixth, there was a small discrepancy in Study C between the self-reported chronic pain diagnosis and the clinic letter diagnosis in Study A1. This could be due to misreporting or evolving diagnoses. Finally, the clinical prospective investigation is limited by its low response rate, small sample size, and the fact that no mothers classified in the other chronic pain group had an autistic child. As a result, the confidence intervals in some of the statistical analysis models are wide, suggesting a high degree of uncertainty. To identify the magnitude and specificity of the association between FMS in mothers and children on the autism spectrum, future research with larger samples or population cohort data should be utilised to improve understanding of the effect size.

For the first time, the present study demonstrated an increased likelihood of autistic children of mothers with FMS (and CRPS) compared to mothers with other chronic pain conditions across multiple, independent cohorts. We suggest asking mothers with FMS and CRPS about autistic children in clinical practice. This may be a valuable addition to clinical routine, allowing better understanding of their home and life situation. More research is required to quantify this correlation and to understand its cause.

## Data Availability

All data produced in the present study are available upon reasonable request to the authors.

