## Supplementary Materials for "INCREASED PREVALENCE OF AUTISM IN THE CHILDREN OF FIBROMYALGIA PATIENTS RELATIVE TO OTHER TYPES OF CHRONIC PAIN"

### Appendix A: *Clinical service evaluation (Study A1)*

#### Supplementary Table 1.

*Chronic pain conditions of patients categorised as "other chronic pain" in the clinical service evaluation (Study A1).*

| "Other" chronic pain condition | N [%] | N of patients with confirmed biological children [%] | N of female patients with confirmed biological children [% of pts with children] | N of female patients with confirmed biological children [% of all pts in category] |
|---|---|---|---|---|
| *Widespread pain* | 134 [33.3%] | 70 [52.2%] | 51 [72.9%] | 51 [38.1%] |
| *Lower body pain* | 82 [20.3%] | 55 [67.1%] | 32 [58.2%] | 32 [39%] |
| *Back pain* | 56 [13.9%] | 39 [70.0%] | 25 [64.1%] | 25 [45%] |
| *Upper body pain* | 20 [5.0%] | 15 [75.0%] | 7 [46.7%] | 7 [35%] |
| *Chronic primary pain* | 14 [3.5%] | 8 [57.1%] | 2 [25%] | 2 [14.3%] |
| *Leg pain* | 9 [2.2%] | 6 [67.0%] | 3 [50%] | 3 [33.3%] |
| *Pelvic pain* | 9 [2.2%] | 2 [22.2%] | 2 [100%] | 2 [22.2%] |
| *Facial pain* | 5 [1.2%] | 4 [80.0%] | 2 [50%] | 2 [40%] |
| *Foot pain* | 5 [1.2%] | 2 [40.0%] | 2 [100%] | 2 [40%] |
| *Knee pain* | 5 [1.2%] | 2 [40.0%] | 2 [100%] | 2 [40%] |
| *Back and leg pain* | 4 [1.0%] | 2 [50.0%] | 1 [50%] | 1 [25%] |
| *Primary chronic pain* | 4 [1.0%] | 4 [100.0%] | 2 [50%] | 2 [50%] |
| *Hip pain* | 3 [0.7%] | 2 [67.0%] | 2 [100%] | 2 [67%] |
| *Leg and foot pain* | 3 [0.7%] | 0 [0.0%] | 0 [0.0%] | 0 [0.0%] |
| *Limb pain* | 3 [0.7%] | 0 [0.0%] | 0 [0.0%] | 0 [0.0% |
| *Neurological disorder* | 3 [0.7%] | 1 [33.3%] | 1 [100%] | 1 [33.3%] |
| *Other* | 44 [11.0%] | 27 [61.4%] | 14 [51.9%] | 14 [31.8%] |

### *Exploratory phenotyping study (Study A2)*

#### Methods

##### *Participants*

FMS patients were recruited to a phenotyping study aiming to correlate clinical with immunological phenotypes, of which, select data from 51 patients has been presented previously [6]. Fifty-nine female FMS patients were included in the current study. Subjects were identified from a registry of patients at a pain management program at the Walton Centre NHS Foundation Trust, for which they had consented for their names to be entered in.

The inclusion criteria required patients with an FMS diagnosis of ACR 2010 [11] or 1990 [12] with a duration of over one year, aged above 18 years, fluent in English, and an average weekly pain intensity >4/10 on a numerical rating scale. The exclusion criteria included participants who were currently pregnancy or breastfeeding.

All participants provided written consent and were reimbursed for up to £30 for their travel expenses. The study received ethical approval from the Health and Care Research Board Wales (Ref: 18/WA/0234).

***Materials***

The study included several validated scales to measure impact and pain indices.

*Impact scales:*

1. Brief Pain Inventory (BPI [10]) is a self-report measure that asks patients to rate their current pain intensity and pain in the last 24 hours at its worst, least, and average on an 11-point numerical scale from *0 (no pain)* to *10 (pain as bad as you can imagine).* Respondents are also asked to rate the extent to which their pain interferes with 7 quality-of-life domains including: general activity, mood, walking ability, normal work (including home and housework), relations with other people, sleep, and enjoyment of life. These are rated on a similar 11-point numerical scale from *0 (does not interfere)* to *10 (completely interferes).* The scale had good internal reliability (McDonald's $\omega_t$ = 0.82).
2. Fibromyalgia Impact Questionnaire-Revised (FIQR [1]) is a 21-item measure, with all questions framed in the context of the past 7 days. There are 3 domains: function,

overall impact, and symptoms. Each item is answered on an 11-point numerical scale from *0* to *10*, with 10 being the 'worst'. The overall scale had excellent internal reliability (McDonald's $\omega_t$ = 0.96).

3. Hospital Anxiety Depression Scale (HADS [8]) is a 14-item measure with 2 subscales, Anxiety and Depression, each with 7 items. Items are scored from *0* to *3*. The overall scale had questionable internal reliability (McDonald's $\omega_h$ = 0.66), however the subscales Anxiety (McDonald's $\omega_t$ = 0.81) and Depression (McDonald's $\omega_t$ = 0.88) displayed good reliability.
4. Pain Catastrophising Scale (PCS [9]) is a 13-item measure of catastrophising. It has 3 subscales; rumination, magnification, and helplessness. All items are referred to a past painful experience and are scored on a 5-point Likert scale ranging from *0 (not at all)* to *4 (all the time)*. The overall scale had acceptable internal reliability (McDonald's $\omega_h$ = 0.72).
5. Pain Self-Efficacy Questionnaire (PSEQ [7]) is a 10-item questionnaire developed to measure the confidence those with ongoing pain have in performing activities while in pain. All items are scored on a 7-point Likert scale ranging from *0 (not at all confident)* to *10 (completely confident)*. The overall scale had excellent internal reliability (McDonald's $\omega_t$ = 0.96).

*Pain scales:*

1. Short Form McGill Pain Questionnaire (SF-MPQ-2 [3]) is a self-report measure consisting of 22 pain descriptors referencing pain and related symptoms in the past week. Items are scored on an 11-point numerical rating scale from *0 (none)* to *10 (worst possible).* Scores are grouped into 4 composite-categories continuous, intermittent, neuropathic, and affective. The overall scale had acceptable internal reliability (McDonald's $\omega_h$ = 0.74), with composite-categories continuous

(McDonald's $\omega_t$ = 0.89), intermittent (McDonald's $\omega_t$ = 0.95), neuropathic (McDonald's $\omega_t$ = 0.92), and affective (McDonald's $\omega_t$ = 0.85) showing good reliability.

***Procedure***

The procedure has been previously described [6]. After providing consent and confirming eligibility, patients were asked questions relating to their general health and fibromyalgia symptomology. This included the question *"Are your children affected by any condition or disorder (including chronic pain)?"* and, if yes, to specify. Additionally, participants completed several standardised questionnaires. The questionnaires included the BPI [10], EQ-5D [5], the FIQR [1], the HADS [8], the SF-MPQ-2 [3], the PCS [9], and the painDETECT [4].

To test mechanical pain threshold and skin sensitivity, a QST protocol was adapted from Boehme et al., [2]. Static pressure pain threshold (PPT) was measured using a 1cm$^2$ rubber-tipped pressure algometer (FDN200; Wagner Instruments) positioned over the vastus lateralis on the left leg. Pressure was increased at a rate of approximately 0.5kg/s (50kPa/s) until the patient stated that the sensation shifted from pressure to pain. Patients were unable to see the pressure dial during testing. Three consecutive readings were taken, and the mean value was used as the PPT score.

Dynamic mechanical allodynia was assessed using brushstroke tests with a QST brush (SENSELab Brush-05, Somedic SenseLab AB, Norra Mellby, Sweden). Light brush strokes were applied manually along a 10 cm segment of non-glabrous skin on the supinated left forearm, moving from proximal to distal in the direction of the hair growth. Each patient received three strokes at a slow (3cm/s) and fast (30cm/s) speed. The speed of the stimulus (slow/fast) was applied alternatively with an interstimulus interval of approximately 30 seconds. Patients were asked to rate the pleasantness of the skin sensation of the brushstroke examination on a grounded 5cm numerical rating scale ranging from *-5 (very unpleasant)* to *+5 (very pleasant).*

**Supplementary Table 2.**

*Patient characteristics of the total sample of female FMS patients. Values represent the median (interquartile range) (Study A2).*

| Patient characteristics | Mothers with non-autistic children | Mothers with autistic children | Females without children | p-value (η2) | Post-Hoc (unadjusted) |
|---|---|---|---|---|---|
| **Demographics** | | | | | |
| *N* | 39 | 5 | 15 | | |
| *Age (yrs)* | 53.00 (8.50) | 49.00 (7.00) | 38.00 (14.00) | **<.001 (0.26)** | Ψ<.001 |
| *BMI (kg/m²)* | 34.80 (8.03) | 32.00 (11.00) | 35.30 (11.20) | .928 (0.00) | |
| *Years of diagnosis* | 8.70 (10.00) | 4.90 (1.30) | 3.90 (3.45) | **.010 (0.13)** | Ψ .005 |
| *Years of WSP symptoms* | 13.10 (12.55) | 8.60 (2.30) | 9.30 (6.85) | .130 (0.04) | |
| **Impact indices** | | | | | |
| *BPI (0-10)* | 7.9 (2.40) | 7.60 (1.40) | 7.90 (2.10) | .533 (0.00) | |
| *FIQR (0-100)* | 74.67 (18.67) | 73.67 (5.33) | 71.17 (18.42) | .315 (0.01) | |
| *EQ-5D Health (0-100)* | 50.00 (28.75) | 40.00 (20.00) | 40.00 (15.00) | .416 (0.00) | |
| *Lack of Energy (0-10)* | 9.00 (2.00) | 8.00 (1.00) | 9.00 (2.00) | .538 (0.00) | |
| *HADS-A (0-22)* | 12.00 (5.50) | 11.00 (2.00) | 12.00 (6.50) | .417 (0.00) | |
| *HADS-D (0-22)* | 12.00 (5.50) | 13.00 (4.00) | 9.00 (5.50) | .438 (0.00) | |
| *PCS (0-52)* | 24.00 (19.75) | 17.00 (11.00) | 20.00 (15.75) | .112 (0.04) | |
| *PSEQ (0-60)* | 17.50 (16.75) | 22.00 (24.00) | 21.00 (8.00) | .178 (0.03) | |
| *SSS (0-12)* | 10.00 (2.00) | 10.00 (1.00) | 9.00 (2.00) | .142 (0.03) | |
| *WPI (0-19)* | 15.00 (5.00) | 10.00 (2.00) | 14.00 (3.00) | **.027 (0.09)** | * .028 |
| **Pain Indices** | | | | | |
| *Ave Pain Resting (0-10)* | 8.00 (1.00) | 7.00 (1.00) | 7.00 (0.00) | .157 (0.03) | |
| *PainDETECT (0-38)* | 26.00 (4.50) | 25.00 (13.00) | 22.00 (9.00) | .411 (0.00) | |
| *SF-M Ave (0-10)* | 6.20 (3.10) | 4.25 (2.27) | 4.40 (3.40) | **.040 (0.08)** | Ψ .030 |
| *SF-M Neuropathic* | 6.35 (3.70) | 4.55 (2.35) | 3.50 (4.00) | .067 (0.06) | |
| *SF-M Affective (0-10)* | 6.50 (3.75) | 4.65 (1.41) | 4.30 (3.30) | .059 (0.07) | |

| | | | | | | |
|---|---|---|---|---|---|---|
| | *SF-M Intermittent (0-10)* | 5.80 (4.25) | 3.40 (2.02) | 4.00 (4.70) | **.049 (0.07)** | *.033 |
| | *SF-M Continuous (0-10)* | 7.30 (1.75) | 4.90 (1.90) | 6.60 (1.80) | .087 (0.05) | |
| **Sensory** | | | | | | |
| | *PPT left leg (kPA)* | 203.00 (115.00) | 236.60 (44.00) | 233.00 (89.00) | .435 (0.01) | |
| | *Pleasantness Slow (-5 to 5)* | 0.00 (2.40) | 0.70 (0.70) | 0.3 (2.15) | .467 (0.00) | |
| | *Pleasantness Fast (-5 to 5)* | 0.00 (1.75) | 0.70 (1.40) | 0.0 (0.85) | .150 (0.03) | |

Abbreviations: BMI, Body Mass Index; HADS- A, Hospital Anxiety Depression Scale – Anxiety; HADS-D, Hospital Anxiety Depression Scale – Depression; EQ-5D, EuroQol 5 Dimensions; SSS, Symptom Severity Score; SF-M Ave, Short Form McGill Average of all ratings; WSP, Widespread Pain; WPI, Widespread Pain Index.

*Significant difference between mothers with children with autism and mothers without children with autism.

Ψ Significant difference between mothers without children with autism and females without children.

### Appendix B: *Online survey (Study B)*

**Questionnaire**

**Your Details:**

- How old are you?
- What is your sex?
- What is your marital status?
- Are you currently pregnant?
- Do you have a chronic pain condition?
- What is your chronic pain condition? (please specify in the box below)

**You and your family's health**

- Do you have a neurodivergent condition? (yes/no)
- Do you have genetically related first-degree family members (parents, siblings, children) with a neurodivergent condition?
  *Neurodivergent conditions include: autism spectrum disorder (including Asperger's syndrome), ADHD, Sensory Processing Disorder*
- If so, how many first-degree family members have a neurodivergent condition?

**Your Pain**

- What year did you first experience your chronic pain symptoms *(or widespread pain symptoms for FMS respondents)*?
- What year were you diagnosed with your chronic pain condition/FMS?
- Who diagnosed you with chronic pain/FMS?
  - (Rheumatologist, GP, pain specialist, other)
- What was your average pain intensity over the past week with *0 = no pain* and *10 = very severe pain?*
- What was the average intensity of fatigue over the past week with *0 = no fatigue* and *10 = very severe fatigue?*

**Your Stress**

- *Every person undergoes stressful events in their lifetime. Lifetime stress refers to the total amount of difficult and stressful life events that a person has experienced over his or her lifespan.* Compared to the average person, would you rate your lifetime stress as:
  - Extremely below average; considerably less than average; slightly less that average ; about average; slightly more than average ; Much more than average; Extremely above average;

**<u>Questionnaire (continued)</u>**

**Your Child, Your Pregnancy and Your Pain During Pregnancy**

- Do you have a child with a neurodivergent condition? If yes, please state what neurodivergent condition.
- *Neurodivergent conditions include: autism spectrum disorder (including Asperger's syndrome), ADHD, Sensory Processing Disorder.*
- If the answer is yes, please state how many of your children are affected with a neurodivergent condition.

(These questions will only be shown if the participants states they have a neurodiverse child)
**THESE QUESTIONS ARE OPTIONAL.**

For each child affected, please answer the following questions. However, if you do not wish to answer, then please exit the survey now.

- How old is your child now? (in years)
- Did your child's other parent have a chronic pain condition at conception/during pregnancy?
- Did you experience the following problems during pregnancy/ birth:
- Gestational age less than 35 weeks
- Low birth weight
- Birth injury or trauma
- Birth defects associated with the central nervous system (e.g. Cerebral palsy)
- Meconium aspiration syndrome
- Neonatal or epileptic encephalopathy
- Genetic disorder (e.g. Down's syndrome)

If you can recall, the following questions relate to your experience around pregnancy with this child.

- If you had **chronic pain/widespread pain before** this pregnancy, how would you rate the intensity of your pain before pregnancy with *0= no pain* and *10 = very severe pain*?

- How would you rate your intensity of your **chronic pain/widespread pain during** pregnancy with *0= no pain* and *10 = very severe pain*?

- How would you rate your level of **fatigue before** this pregnancy with *0= no fatigue* and *10 = very severe fatigue*?

- How would you rate your intensity of **fatigue during** this pregnancy with *0= no fatigue* and *10 = very severe fatigue*?

**Supplementary Table 3.**

*Most reported chronic pain conditions among the "other chronic pain" group (Study B).*

| "Other" chronic pain condition | N [%] |
|---|---|
| *Back pain* | 26 [21.49%] |
| *Arthritis* [a] | 11 [9.09%] |
| *Osteoarthritis* | 7 [5.79%] |
| *Neck/shoulder pain* | 7 [5.79%] |
| *Headaches/migraines* | 6 [4.96%] |
| *Joint pain* | 6 [4.96%] |
| *Sciatica* | 6 [4.96%] |
| *Pelvic pain* | 4 [3.31%] |
| *Chronic pain syndrome* | 3 [2.48%] |
| *Facial pain* | 3 [2.48%] |
| *Muscle pain* | 3 [2.48%] |
| *Neuropathic pain* | 3 [2.48%] |
| *Abdominal pain* | 2 [1.65%] |
| *Back and other localised pain* | 2 [1.65%] |
| *Back pain and arthritis* | 2 [1.65%] |
| *Diabetes related pain* | 2 [1.65%] |
| *Ehlers-Danlos syndrome* | 2 [1.65%] |
| *Nerve damage* | 2 [1.65%] |
| *Pain from autoimmune condition* | 2 [1.65%] |
| *Other* | 21 [17.36%] |

[a] These respondents did not specify the type of arthritis.
*Note.* Conditions included in "other" count were each only named once.

**Supplementary Table 4.**

*Most reported diagnosed neurodivergent conditions among offspring, stratified by maternal chronic pain condition (Study B).*

| | Mothers with FMS | Mothers with other chronic pain |
|---|---|---|

| Most reported diagnosed neurodivergent conditions in offspring | N [%] Total including co-occurring conditions | N Mothers with only one neurodiverse child and with only a single neurodiverse condition stated | N [%] Total including co-occurring conditions | N Mothers with only one neurodiverse child and with only a single neurodiverse condition stated |
|---|---|---|---|---|
| *N* | 528 | | 94 | |
| *Autism* | 137 [25.95%] | | 10 [10.64%] | |
| *Attention Deficit Hyperactivity Disorder* | 115 [21.78%] | 64 [12.12%] | 14 [14.89%] | 8 [8.51%] |
| *Sensory processing disorder* | 23 [4.36%] | 19 [3.60%] | 2 [2.13%] | 1 [1.06%] |
| *Learning disability* | 11 [2.08%] | 9 [1.70%] | 0 [0.00%] | 0 [0.00%] |
| *Obsessive Compulsive Disorder (OCD)* | 9 [1.70%] | 9 [1.70%] | 1 [1.06%] | 0 [0.00%] |
| *Dyslexia* | 4 [0.76%] | 3 [0.57%] | 0 [0.00%] | 0 [0.00%] |
| *Dyspraxia* | 4 [0.76%] | 2 [0.38%] | 1 [1.06%] | 0 [0.00%] |
| *Tourette's syndrome* | 3 [0.57%] | 1 [0.19%] | 0 [0.00%] | 0 [0.00%] |
| *Semantic Pragmatic Disorder* | 2 [0.38%] | 0 [0.00%] | 0 [0.00%] | 0 [0.00%] |
| *Oppositional Defiant Disorder (ODD)* | 2 [0.38%] | 0 [0.00%] | 1 [1.06%] | 0 [0.00%] |

*Note.* Values in the total column include multiple entries per respondent due to co-occurring conditions in the same child, and as mothers may have had more than one neurodiverse child (see main text methods section).

### Supplementary Table 5.

*Participant characteristics of the fathers-only and the mothers-only sample (Study B).*

| | Fathers-only sample | | Mothers-only sample | |
|---|---|---|---|---|
| | FMS | Other chronic pain [a] | FMS | Other chronic pain [a] |
| **Characteristics** | N [%] or M (SD) | N [%] or M (SD) | N [%] or M (SD) | N [%] or M (SD) |
| *N* | 57 | 23 | 528 | 94 |
| *Age yrs* | 31.89 (7.93) | 38.00 (12.69) | 42.00 (11.32) | 45.14 (16.09) |
| *Respondents with a diagnosed neurodivergent condition* | 29 [50.88%] | 16 [69.57%] | 149 [28.21%] | 41 [43.62%] |

| | | | | |
|---|---|---|---|---|
| *1st degree family with diagnosed neurodivergent condition* [c] | 21 [36.84%] | 15 [65.21%] | 110 [20.83%] | 31 [32.98%] |
| *Weekly average pain score (0-10)* | 5.96 (1.91) | 4.78 (1.65) | 6.71 (1.75) | 5.45 (1.98) |
| *High lifetime-stress estimate (much more/extremely above average)* | 9 [15.79%] | 3 [13.04%] | 320 [60.61%] | 31 [32.98%] |
| ***At least one autistic child*** | **3 [5.25%]** | **1 [4.35%]** | **137 [25.95%]** | **10 [10.64%]** |
| *Answered "Yes" to having a neurodiverse child* | 13 [22.81%] | 2 [8.70%] | 304 [57.58%] | 25 [26.60%] |
| *Child with ADHD* [b] | 6 [10.53%] | 1 [4.35%] | 115 [21.78%] | 14 [14.89%] |

[b] Some may represent co-occurring ADHD with autism or other ND condition.
[c] First-degree family include the respondent's parents and siblings, but not children.

**Supplementary Table 6.**

*Characteristics of mothers with FMS and other chronic pain, stratified by the ADHD status of children (Study B).*

| | Mothers with FMS | | Mothers with other chronic pain | |
|---|---|---|---|---|
| | **At least one ADHD child** | **Non-ADHD children** | **At least one ADHD child** | **Non-ADHD children** |
| **Characteristics** | N [%] or M (SD) | N [%] or M (SD) | N [%] or M (SD) | N [%] or M (SD) |
| *N* | 115 [21.78%] | 413 [78.22%] | 14 [14.89%] | 80 [85.11%] |
| *Age yrs* | 41.7 (10.70) | 42.1 (11.50) | 46.1 (13.30) | 45.0 (16.6) |
| *Respondents with a diagnosed neurodivergent condition* | 42 [36.52%] | 107 [25.91%] | 8 [57.14%] | 33 [41.25%] |
| *1st degree family with diagnosed neurodivergent condition* [a] | 24 [20.87%] | 86 [20.82%] | 3 [21.43%] | 28 [35.00%] |
| *Weekly average pain score (0-10)* | 7.25 (1.52) | 6.56 (1.79) | 6.00 (2.57) | 5.35 (1.86) |
| *High lifetime-stress estimate (much* | 79 [68.69%] | 241 [58.35%] | 5 [35.71%] | 26 [32.50%] |

*more/extremely above average)*

[a] First-degree family include the respondent's parents and siblings, but not children.
*Note.* Some mothers reported multiple neurodiverse children and the same child with ADHD may have a co-occurring neurodivergent condition (e.g., autism). Maternal chronic pain condition was not significantly associated with ADHD in offspring (OR= 1.55, 95% CIs 0.88 – 2.92, $p$=.134).

**Supplementary Figure 1.**

*Box plot showing the distribution of maternal age at the birth of children with an autism diagnosis. Online survey (Study B).*

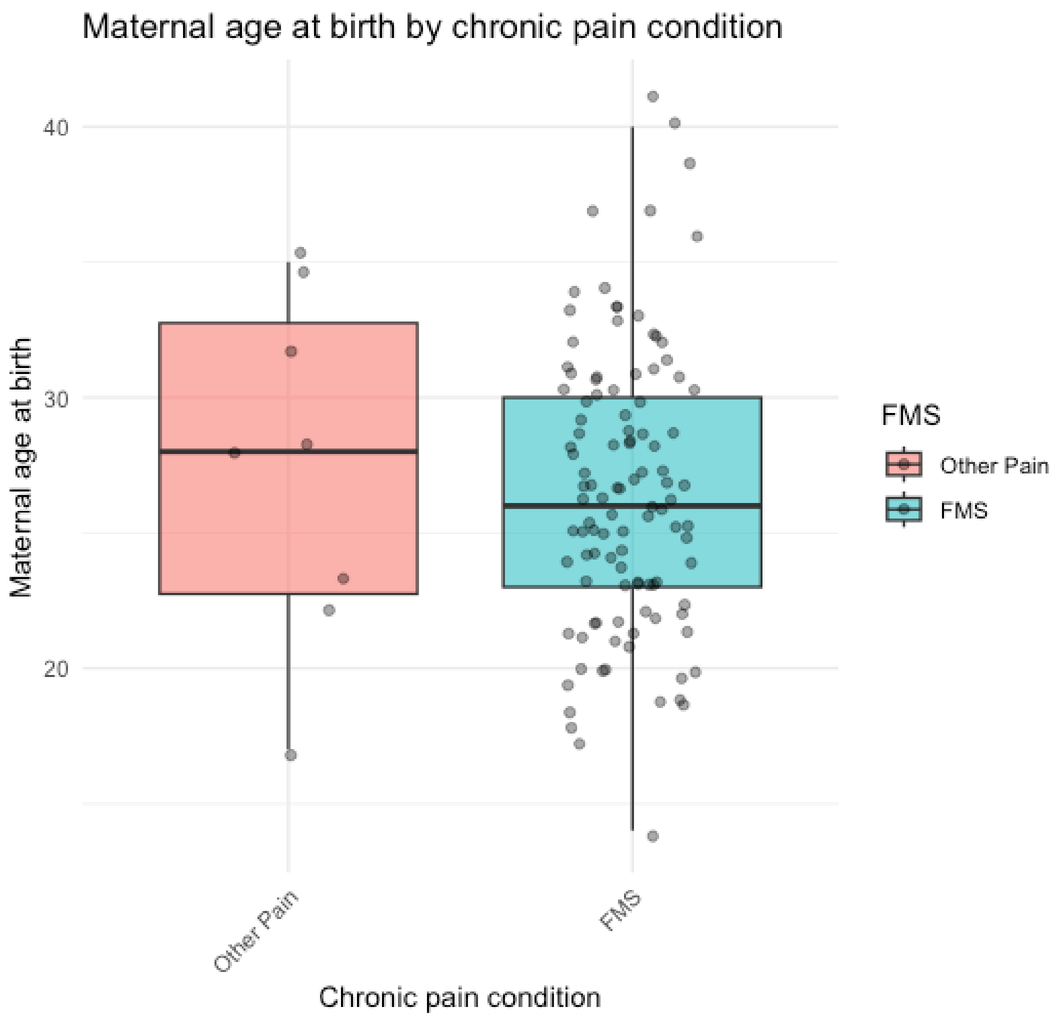

**Supplementary Table 7.**

*Penalised logistic regression analysis showing the association of parental sex to children with an autism diagnosis.*

| Predictors | Model 1: | | | Model 2: | | |
|---|---|---|---|---|---|---|
| | B (SE) | OR (95% CIs) | *p* value | B (SE) | OR (95% CIs) | *p* value |
| Model 1 | | | | | | |
| *Parental sex (mothers vs. fathers)* | 1.57 (0.49) | 4.82 (2.03 – 14.85) | **<.001** | 0.62 (0.90) | 0.96 (0.40 – 17.91) | .463 |
| *Parental chronic pain condition (FMS vs. Other chronic pain)* | 0.99 (0.33) | 2.68 (1.47 – 5.35) | **<001** | -0.04 (1.01) | 1.86 (0.15 – 10.35) | .970 |
| Model 2: Interaction | | | | | | |
| *Parental sex * Parental chronic pain condition* | **-** | **-** | **-** | 1.07 (1.06) | 2.93 (0.25 – 21.54) | .341 |

**Supplementary Table 8.**

*Penalised logistic regression analysis showing the association of a high self-reported lifetime stress estimate to children with an autism diagnosis.*

| Maternal predictors | Model 1: | | | Model 2: | | |
|---|---|---|---|---|---|---|
| | B (SE) | OR (95% CIs) | *p* value | B (SE) | OR (95% CIs) | *p* value |
| Model 1 | | | | | | |
| *High lifetime stress estimate (much more/extremely above average)* | 0.41 (0.20) | 1.51 (1.02 - 2.24) | **.037** | 0.37 (0.66) | 1.45 (0.37 – 5.19) | .576 |
| *Maternal chronic pain conditions (FMS vs. Other pain)* | 0.93 (0.35) | 2.53 (1.34 – 5.27) | **.003** | 0.87 (0.45) | 2.39 (1.06 – 6.28) | **.035** |
| Model 2: Interaction | | | | | | |
| *High lifetime stress * Maternal chronic pain condition* | **-** | **-** | **-** | 0.05 (0.69) | 1.05 (0.27 – 4.31) | .944 |

**Appendix C:** ***Clinical prospective investigation (Study C)***

**Questionnaire**

**Your details**

1. How old are you?
2. What is your sex?
3. What is your marital status?
4. Are you currently pregnant? If yes, exited out of questionnaire.
5. What is your diagnosed chronic pain condition/ where is your chronic pain located?
6. What year were you diagnosed with this chronic pain condition?
7. Before the development of your chronic pain condition, did you have any other chronic pain conditions? If yes, when did this start?

**GAD- 7: Over the last 2 weeks, how often have you been bothered by any of the following problems?**

1. Feeling nervous, anxious, or on edge?
2. Not being able to stop or control worrying?
3. Worrying too much about different things?
4. Trouble relaxing?
5. Being so restless that it is hard to sit still?
6. Becoming easily annoyed or irritable?
7. Feeling afraid as if something awful might happen?

**AQ-10: Please respond with the answer that most accurately describes how each of the statements below applies to you.**

1. I often notice small sounds when others do not.
2. When I'm reading a story, I find it difficult to work out the characters' intentions.
3. I find it easy to "read between the lines" when someone is talking to me.
4. I usually concentrate more on the whole picture, rather than the small details.
5. I know how to tell if someone listening to me is getting bored.
6. I find it easy to do more than one thing at once.
7. I find it easy to work out what someone is thinking or feeling just by looking at their face.

**Questionnaire (continued)**

8. If there is an interruption, I can switch back to what I was doing very quickly.

9. I like to collect information about categories of things.

10. I find it difficult to work out people's intentions.

**RAADS-14: Please respond with the answer that most accurately describes how each of the statements below applies to you. For the purposes of this test, "*When I was Young*" refers to the age of 16 or younger.**

1. It is difficult for me to understand how other people are feeling when they are talking.

2. Some ordinary textures that do not bother others feel very offensive when they touch my skin.

3. It is very difficult for me to work and function in groups.

4. It is difficult to figure out what other people expect of me.

5. I often don't know how to act in social situations.

6. I can chat and make small talk with people. *

7. When I feel overwhelmed by my senses I have to isolate myself to shut them down.

8. How to make friends and socialise is a mystery to me.

9. When talking to someone, I have a hard time telling when it is my turn to talk or to listen.

10. Sometimes I have to cover my ears to block out painful noises (like vacuum cleaners or people talking too much or too loudly)

11. It can be very hard to read someone's face, hand, and body movements when they are talking.

12. I focus on the details rather than overall idea

13. I take things too literally, so I often miss what people are trying to say.

14. I get extremely upset when the way I like to do things is suddenly changed.

**Family neurodivergence**

1. Have you been diagnosed with a neurodivergent condition?
   ***Neurodivergent conditions include: Autism Spectrum Disorder (ASD), Attention Deficit Hyperactivity Disorder (ADHD), Sensory processing disorder, dyspraxia.***
   - If yes, what condition?
   - Which medical professional diagnosed you with this neurodivergent condition?

**Questionnaire (continued)**

2. Do you have genetically related first-degree family members with a diagnosed neurodivergent condition?
   ***First-degree relatives include an individual's parents, siblings and offspring.***
   - If yes, who has been affected?
     a) Parent(s)
     b) Sibling(s)
     c) Child(ren)
   - What condition do they have?

3. Do you have children?

**Your pain, pregnancy, and children. Please complete these questions in the birth order of all your children, starting with your firstborn (oldest) and your experience during this pregnancy.**

1. Did you have chronic pain at the time of this pregnancy, or in the year following this pregnancy? If yes, where was this located and what year did this start?

2. If you had chronic pain during pregnancy/in the year following, how would you rate the severity of this pain at the time of pregnancy?

3. If you can remember the severity of this pain, did the intensity of this pain change during pregnancy? (increase, decrease, or stay the same).

4. Did you smoke during pregnancy?

5. What is this child's sex?

6. How old is this child now?

**Parent-report AQ-10:**

1. S/he often notices small sounds when others do not.

2. S/he usually concentrates more on the whole picture, rather than the small details.

3. In a social group, s/he can easily keep track of several different people's conversations.

4. S/he finds it easy to go back and forth between activities.

5. S/he doesn't know how to keep a conversation going with his/her peers

6. S/he is good at social chit-chat

7. When s/he is reading a story, s/he finds it difficult to work out the character's intentions or feelings.

8. When s/he was in preschool, s/he used to enjoy playing games involving pretending with other children.

**<u>Questionnaire (continued)</u>**

9. S/he finds it easy to work out what someone is thinking or feeling just by looking at their face.

10. S/he finds it hard to make new friends.

**The next set of questions still refer to your first-born (oldest) child.**

1. Does your first-born (oldest) child have an autism spectrum disorder diagnosis?
- If yes, which medical professional diagnosed your child with autism spectrum disorder?
- How old was your child at the time of diagnosis?

2. If no, have you had concerns about your first-born (oldest) child's social or communication development, or their rigid/repetitive interests or behaviours?

**SCQ:**

1. Is s/he now able to talk using short phrases or sentences? **If *no* please skip to question 8.**

2. Can you have a to-and-fro "conversation" with her/him that involves taking turns or building on what you have said?

3. Has s/he ever used odd phrases or said the same thing over and over in almost exactly the same way (either phrases that s/he hears other people use of ones that s/he makes up)?

4. Has s/he ever used socially inappropriate questions or statements? For example, has s/he ever regularly asked personal questions or made personal comments at awkward times?

5. Has s/he ever got her/his pronouns mixed up? For example, saying *you* or *she/he* for *I*?

6. Has s/he ever used words that s/he seemed to have invented or made up her/himself; put things in odd, indirect ways; or use metaphorical ways of saying things? For example, saying *hot rain* for *steam?*

7. Has s/he ever said the same thing over and over in exactly the same way or insist that you say the same thing over and over again?

8. Has s/he ever had things that s/he seems to have to do in a very particular way or order or rituals that s/he insists that you go through?

9. Has her/his facial expression usually seemed appropriate to the particular situation, as far as you can tell?

10. Has s/he ever used your hand like a tool or as if it were part of her/his own body? For example, pointing with your finger or putting your hand on a doorknob to get you to open the door?

11. Has s/he ever had any interests that preoccupy her/him and might seem odd to other people? For example, traffic lights, drainpipes, timetables.

**Questionnaire (continued)**

12. Has s/he ever seemed to be more interested in parts of a toy or an object, rather than in using the object as it was intended? For example, spinning the wheels of a toy car.

13. Has s/he ever had any special interests that are unusual in their intensity but otherwise appropriate for her/his age and peer group? For example, trains or dinosaurs

14. Has s/he ever seemed to be unusually interested in sight, feel, sound, taste, or smell of things or things or people?

15. Has s/he ever had any mannerisms or odd ways of moving her/his hands or fingers, such as flapping or moving her/his fingers in front of her/his eyes?

16. Has s/he ever had any complicated movements of her/his whole body, such as spinning or repeatedly bouncing up and down?

17. Has s/he ever injured her/himself deliberately, such as by biting her/his arm or banging her/his head?

18. Has s/he ever had any objects (other than a soft toy or comfort blanket) that s/he has to carry around?

19. Does s/he have any particular friends or a best friend?

20. When s/he was 4 to 5, did s/he ever talk with you just to be friendly (rather than to get something)?

21. When s/he was 4 to 5, did s/he ever spontaneously copy you (or other people) or what you are doing (such as vacuuming, gardening, or mending things)?

22. When s/he was 4 to 5, did s/he ever spontaneously point at things around her/him just to show you things (not because s/he wants them)?

23. When s/he was 4 to 5, did s/he ever use gestures, other than pointing or pulling your hand, to let you know what s/he wants?

24. When s/he was 4 to 5, did s/he nod her/his head to indicate yes?

25. When s/he was 4 to 5, did s/he shake her/his head to indicate no?

26. When s/he was 4 to 5, did s/he usually look at you directly in the face when doing things with you or talking with you?

27. When s/he was 4 to 5, did s/he smile back if someone smiles at her/him?

28. When s/he was 4 to 5, did s/he ever show you things that interest her/him to engage your attention?

29. When s/he was 4 to 5, did s/he ever offer to share things other than food with you?

**Questionnaire (continued)**

30. When s/he was 4 to 5, did s/he ever seem to want you to join in her/his enjoyment of something?
31. When s/he was 4 to 5, did s/he ever try to comfort you if you were sad or hurt?
32. When s/he was 4 to 5, when s/he wanted something or wanted help, did s/he look at you and use gestures with sounds or words to get your attention?
33. When s/he was 4 to 5, did s/he show a normal range of facial expression?
34. When s/he was 4 to 5, did s/he ever spontaneously join in and try to copy the actions in social games, such as *The Mulberry Bush* or *London Bridge Is Falling Down?*
35. When s/he was 4 to 5, did s/he play any pretend or make-believe games?
36. When s/he was 4 to 5, did s/he seem interested in other children of approximately the same age whom s/he did not know?
37. When s/he was 4 to 5, did s/he respond positively when another child approached her/him?
38. When s/he was 4 to 5, if you came into a room and started talking to her/him without calling her/his name, did s/he usually look up and pay attention to you?
39. When s/he was 4 to 5, did s/he ever play imaginative games with another child in such a way that you can tell that each child understood what the other was pretending?
40. When s/he was 4 to 5, did s/he play cooperatively in games that need some form of joining in with a group of other children, such as hide-and-seek or ball games?

**If your first-born (oldest) child has an autism spectrum disorder diagnosis, we would like to ask you some questions about this child's health and autism spectrum disorder.**

1. Does your child have any other medical diagnoses, including genetic or medical conditions?
2. Does your child have an intellectual disability?
   - If yes, and you know the "severity" of your child's intellectual disability, how would it best be described? (Mild, Moderate, Severe/profound).
3. Did your child have any complications during pregnancy, delivery, or after their birth?
4. When your child was diagnosed with autism, the doctor may have assigned a level 1, 2, or 3 to include two areas of functioning: a) social communication b) restricted, repetitive, behaviours. Do you know the level of support needs for your child?
   - If yes, what is the level of support specified for your child?
5. Is there anything else you would like to tell us about your child's health, or about anything else that we have asked in this survey?
6. Do you have another child?

**Questions are repeated for each child.**

**Supplementary Table 9.**

*Patient characteristics of the total sample and the mothers-only sample by chronic pain condition (Study C).*

| | Total sample | | | Mothers-only sample | | |
|---|---|---|---|---|---|---|
| | **FMS** | **CRPS** | **Other chronic pain** | **FMS** | **CRPS** | **Other chronic pain** |
| **Patient characteristics** | N [%] or median (IQR) | N [%] or median (IQR) | N [%] or median (IQR) | N [%] or median (IQR) | N [%] or median (IQR) | N [%] or median (IQR) |
| *N* | 29 | 11 | 26 | 24 | 7 | 11 |
| *Age (yrs) (mean (SD))* | 45.8 (1.17) | 48.3 (1.45) | 51.0 (1.10) | 49.1 (8.28) | 54.1 (9.87) | 49 (14.20) |
| *Sex = female* | 27 [93.10%] | 8 [72.73%] | 15 [57.69%] | - | - | - |
| *Biological children* | 26 [89.66%] | 9 [81.82%] | 22 [84.62%] | - | - | - |
| ***Autistic child*** | **8 [27.59%]** | **3 [27.27%]** | **3 [11.54%]** | **8 [33.33%]** | **2 [28.57%]** | **0 [0.00%]** |
| *Autistic respondent* | 3 [10.34%] | 1 [9.09%] | 1 [3.85%] | 2 [8.33%] | 0 [0.00%] | 0 [0.00%] |
| *1st degree family with diagnosed neurodivergent condition [a]* | 5 [17.24%] | 3 [27.27%] | 4 [15.38%] | 5 [20.83%] | 1 [14.29%] | 2 [18.18%] |
| *GAD-7 (0-21)* | 7 (9) | 9 (11) | 10.5 (10.8) | 7.5 (9.5) | 4 (11) | 13 (8.5) |
| *Scored above cutoff (≥10* | 12 [41.38%] | 5 [45.45%] | 14 [53.85%] | 10 [41.67%] | 3 [42.86%] | 7 [63.64%] |
| *RAADS-14 Total (0-42)* | 27 (21) | 23 (14) | 21.5 (23.5) | 22 (20) | 20 (22) | 24 (16) |
| *Scored above cutoff (≥14)* | 21 [72.41%] | 8 [72.73%] | 19 [73.08%] | 16 [66.67%] | 5 [71.43%] | 9 [81.82%] |
| *AQ-10 (0-10, cutoff ≥6)* | 4 (1) | 5 (2) | 5 (2) | 4.5 (1.25) | 5 (1.5) | 5 (1.5) |
| *Scored above cutoff (≥6)* | 7 [24.14%] | 5 [45.45%] | 8 [30.77%] | 6 [25.00%] | 3 [42.86%] | 3 [27.27%] |

[a] First-degree family include the respondent's parents and siblings, but not children.
Abbreviations: IQR, Interquartile Range. SD, Standard Deviation.

**Supplementary Table 10.**

*Characteristics of mothers with FMS, CRPS and other chronic pain, stratified by the autism status of children (Study C).*

| | Mothers with FMS | | Mothers with CRPS | | Mothers with other chronic pain |
|---|---|---|---|---|---|
| | **Autistic child** | **Non-autistic child** | **Autistic child** | **Non-autistic child** | **Non-autistic child** |
| **Patient characteristics** | N [%] or median (IQR) | N [%] or median (IQR) | N [%] or median (IQR) | N [%] or median (IQR) | N [%] or median (IQR) |
| *N* | 8 | 16 | 2 | 5 | 11 |
| *Age (yrs) (mean (SD)* | 48.8 (7.34) | 49.3 (8.93) | 40.5 (0.71) | 59.6 (3.97) | 49 (14.2) |
| *Autistic respondent* | 0 [0.00%] | 2 [12.5%] | 0 [0.00%] | 0 [0.00%] | 0 [0.00%] |
| *1st degree family with diagnosed neurodivergent condition* [a] | 3 [37.50%] | 1 [6.25%] | 0 [0.00%] | 1 [20.00%] | 2 [18.18%] |
| *GAD-7 (0-21)* | 7 (8) | 7.5 (11.2) | 16.5 (3.5) | 4 (1) | 13 (8.5) |
| *Scored above cutoff (≥10)* | 3 [37.50%] | 7 [43.75%] | 2 [100.00%] | 1 [20.00%] | 7 [63.64%] |
| *RAADS-14 Total (0-42)* | 25 (14.5) | 21.5 (20) | 40.5 (1.5) | 18 (18) | 24 (16) |
| *Scored above cutoff (≥14)* | 6 [75.00%] | 10 [62.50%] | 2 [100.00%] | 3 [60.00%] | 9 [81.82%] |
| *AQ-10 (0-10)* | 4.5 (1.25) | 4.5 (1.25) | 5.5 (0.5) | 5 (2) | 5 (1.5) |
| *Scored above cutoff (≥6)* | 2 [25.00%] | 4 [25.00%] | 1 [50.00%] | 2 [40.00%] | 3 [27.27%] |

[a] First-degree family include the respondent's parents and siblings, but not children.
Abbreviations: IQR, Interquartile Range. SD, Standard Deviation.

**Supplementary Table 11.**

*Characteristics of autistic children by parental chronic pain condition in the clinical prospective investigation (Study C).*

| | FMS | CRPS | Other chronic pain |
|---|---|---|---|
| **Characteristics of autistic child** | N [%] or mean (SD) | N [%] or mean (SD) | N [%] or mean (SD) |
| *N (of parents with that chronic pain condition)* | 8 | 3 | 3 |
| *Parental age at birth* | 27.12 (6.83) | 28.33 (6.11) | 34.67 (2.52) |
| *AQ-10 score median (IQR)(0-10)* | 8.50 (1.50) | 9.00 (0.50) | 7.00 (2.00) |
| *Scored above cutoff( ≥6)* | 8 [100.00%] | 3 [100.00%] | 2 [66.67%] |
| *SCQ score median (IQR) (0-39)* | 28.00 (6.50) | 31.00 (2.50) | 17.00 (6.00) |
| *Scored above cutoff( ≥15)* | 8 [100.00%] | 3 [100.00%] | 3 [100.00%] |
| *Age at diagnosis yrs* | 14.29 (4.82) | 5.83 (5.48) | 8.67 (4.16) |
| *Sex = male* | 4 [50.00%] | 3 [100.00%] | 2 [66.67%] |
| *Birth order:* | | | |
| *1st child:* | 3 [37.50%] | 2 [66.67%] | 2 [66.67%] |
| *2nd child:* | 2 [25.00%] | - | 1 [33.33%] |
| *3rd child:* | 3 [37.5%] | 1 [33.33%] | - |
| *Any maternal chronic pain during pregnancy* | 4 [50.00%] | 2 [66.67%] | Unknown |
| *Birth/ pregnancy complications* | 2 [25.00%]<br>• Respiratory distress syndrome<br>• Low blood sugar | 2 [66.67%)<br>• Low birth weight<br>• Premature, sepsis and meningitis, necrosis of placenta | - |
| *Co-occurring intellectual disability* | 3 [37.50%] | 2 [66.67%] | 1 [33.33%] |
| *Level of support needs:* | | | |
| *Level 1:* | - | - | Unknown |
| *Level 2:* | 3 [37.50%] | - | - |
| *Level 3:* | - | 2 [66.67%] | - |
| *Child's other co-occurring disorders/ conditions* | • Pulmonary stenosis<br>• Cerebral Palsy<br>• Asthma (x2)<br>• ADHD | • ADHD, global developmental delay, asthma.<br>• Angelman syndrome | • Global delay |

*Note.* Parents did not know or report answers for all of the reported variables.

**Supplementary Table 12.**

*Maternal data gathered from all pregnancies, stratified by maternal chronic pain condition and autism diagnosis status of children (Study C).*

| | Mothers with FMS | | Mothers with CRPS | | Mothers with other chronic pain |
|---|---|---|---|---|---|
| | **Pregnancies of autistic children** | **Pregnancies of non-autistic children** | **Pregnancies of autistic children** | **Pregnancies of non-autistic children** | **Pregnancies of non-autistic children** |
| *Number of pregnancies (and children)* | 8 | 43 | 2 | 11 | 22 |
| *Reported pain during pregnancy* | 4 [50.00%] | 20 [46.51%] | 2 [100.00%] | 3 [27.27%] | 15 [68.18%] |
| | - Widespread pain (x2)<br>- Right leg pain after DVT<br>- Hip pain, SPD, migraines | - FMS and back pain<br>- FMS (x11)<br>- Back pain<br>- Back and neck<br>- Back, hips, legs<br>- Arthritis in back and symphysis (x2)<br>- CRPS (x3) | - CRPS (x2) | - CRPS (x2)<br>- Did not specify | - Joint pain (x2)<br>- Sciatica<br>- Back pain x 8<br>-EDS (x2)<br>- Full body pain<br>- Knee pain |
| *If pain, intensity of pain during pregnancy:* | | | | | |
| *Mild:* | 1 [12.50%] | 0 [0.00%] | 1 [50.00%] | 0 [0.00%] | 1 [4.55%] |
| *Moderate:* | 1 [12.50%] | 8 [18.60%] | 0 [0.00%] | 2 [18.18%] | 6 [27.27%] |
| *Severe:* | 2 [25.00%] | 10 [23.26%] | 1 [50.00%] | 1 [9.09%] | 8 [36.36%] |
| *Unknown*:* | 0 [0.00%] | 2 [4.65%] | 0 [0.00%] | 0 [0.00%] | 0 [0.00%] |
| *If pain, difference in pain intensity from before to during pregnancy:* | | | | | |
| *Increase:* | 2 [25.00%] | 9 [20.93%] | 1 [50.00%] | 1 [9.09%] | 10 [45.45%] |
| *Stayed the same:* | 1 [12.50%] | 3 [6.98%] | 0 [0.00%] | 0 [0.00%] | 5 [22.73%] |
| *Decrease:* | 0 [0.00%] | 4 [9.30%] | 1 [50.00%] | 2 [18.18%] | 0 [0.00%] |
| *Unknown*:* | 1 [12.50%] | 4 [9.30%] | 0 [0.00%] | 0 [0.00%] | 0 [0.00%] |

Abbreviations: CRPS, Complex Regional Pain Syndrome; DVT, Deep Vein Thrombosis; EDS, Ehlers-Danlos Syndrome; FMS, Fibromyalgia Syndrome; SPD, Symphysis Pubis Dysfunction.
*Note.* Reported pain during pregnancy was counted per pregnancy.
*Patients who did not know or remember the intensity or difference in intensity of pain during pregnancy.

**Supplementary Table 14.**

*AQ-10 and SCQ total scores of all children, by parental (mothers and fathers) chronic pain condition and autism diagnosis status. Values represent median (interquartile range). (Study C).*

| | Parent with FMS | | Parent with CRPS | | Parent with other chronic pain | |
|---|---|---|---|---|---|---|
| | **Autistic child** | **Non-autistic child** | **Autistic child** | **Non-autistic child** | **Autistic child** | **Non-autistic child** |
| *N children* | 8 | 45 | 3 | 14 | 3 | 37 |
| *AQ-10 total score (0-10)* | 8.50 (1.50) | 3.00 (5.00) | 9.00 (0.50) | 1.00 (6.00) | 7.00 (2.00) | 4.00 (4.00) |
| *N [%] scored above AQ-10 cutoff(* ≥6) | 8 [100%] | 16 [35.56%] | 3 [100%] | 5 [35.71%] | 2 [66.67%] | 11 [29.73%] |
| *SCQ total score* (0-39; cut off*≥15) | 24.00 (6.50) | 11.50 (12.20) | 31.00 (2.50) | 17.00 (11.00) | 17.00 (6.00) | 16.00 (7.50) |
| *N [%] scored above SCQ cutoff(≥15)* | 8 [100%] | 8 [17.78%] | 3 [100%] | 4 [28.57%] | 3 [100%] | 9 [24.32%] |

*The SCQ was only completed by parents who had concerns about their child's development or had autism. **Regarding non-autistic children, FMS patients reported concern for the development of 20/45 (44.44%) children, CRPS 5/15 (33.33%) children, and other chronic pain 17/37 (45.95%) children.**

**Supplementary Table 15.**

*AQ-10 and SCQ total scores of non-autistic children, by parental (mothers and fathers) chronic pain condition and whether they had an autistic sibling. Values represent median (interquartile range). (Study C).*

| | Parent with FMS | | Parent with CRPS | | Parent with other chronic pain | |
|---|---|---|---|---|---|---|
| | **Has an autistic sibling** | **No autistic siblings** | **Has an autistic sibling** | **No autistic siblings** | **Has an autistic sibling** | **No autistic siblings** |
| *N children* | 13 | 32 | 5 | 10 | 3 | 35 |
| *AQ-10 total score (0-10; cutoff* ≥6) | 3.00 (5.00) | 3.00 (5.25) | 6.00 (7.00) | 1.00 (4.75) | 3.00 (0.50) | 4.50 (4.75) |
| *N [%]scored above AQ-10 cutoff(* ≥6) | 5 [38.46%] | 11 [34.38%] | 3 [60.00%] | 2 [20.00%] | 0 [0.00%] | 11 [31.43%] |

| | | | | | | |
|---|---|---|---|---|---|---|
| *SCQ total score* (0-39; cut off≥15)* | 11.00 (16.00) | 12.00 (9.00) | 24.50 (7.50) | 15.00 (8.50) | NA | 16.00 (7.50) |
| *N [%] scored above SCQ cutoff(≥15)* | 3 [23.08%] | 5 [15.63%] | 2 [40.00%] | 2 [20.00%] | NA | 9 [25.71%] |

*The SCQ was only completed by parents who had concerns about their child's development. **Regarding non-autistic children, FMS patients reported concern for the development of 20/45 (44.44%) children, CRPS 5/15 (33.33%) children, and other chronic pain 17/37 (45.95%) children.**

**Supplementary Figure 2.**

*Age of autism spectrum diagnosis by sex and parental chronic pain condition. Clinical prospective investigation (Study C).*

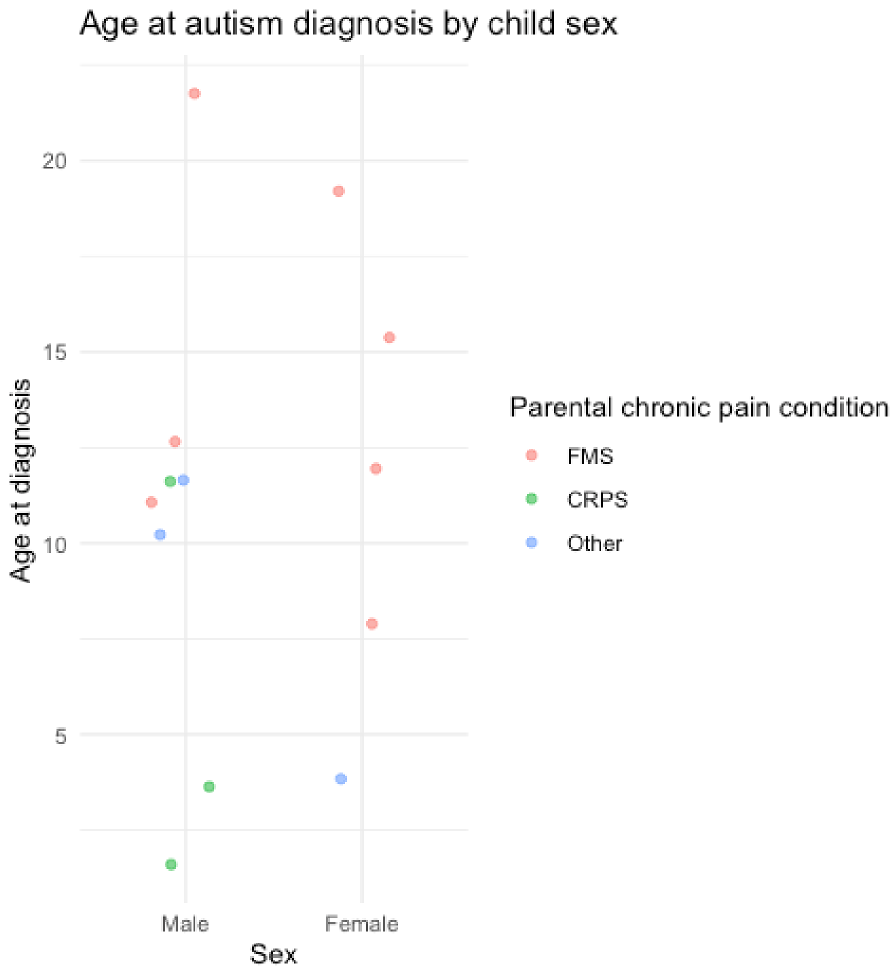

[1] Bennett RM, Friend R, Jones KD, Ward R, Han BK, Ross RL. The revised fibromyalgia impact questionnaire (FIQR): validation and psychometric properties. Arthritis research & therapy 2009;11(4):R120. doi: 10.1186/ar2783
[2] Boehme R, van Ettinger-Veenstra H, Olausson H, Gerdle B, Nagi SS. Anhedonia to gentle touch in fibromyalgia: normal sensory processing but abnormal evaluation. Brain Sciences 2020;10(5):306. doi: 10.3390/brainsci10050306
[3] Dworkin RH, Turk DC, Revicki DA, Harding G, Coyne KS, Peirce-Sandner S, Bhagwat D, Everton D, Burke LB, Cowan P. Development and initial validation of an expanded and revised version of the Short-form McGill Pain Questionnaire (SF-MPQ-2). Pain® 2009;144(1-2):35-42. doi: 10.1016/j.pain.2009.02.007
[4] Freynhagen R, Baron R, Gockel U, Tölle TR. Pain DETECT: a new screening questionnaire to identify neuropathic components in patients with back pain. Current medical research and opinion 2006;22(10):1911-1920. doi: 10.1185/030079906X132488
[5] Herdman M, Gudex C, Lloyd A, Janssen M, Kind P, Parkin D, Bonsel G, Badia X. Development and preliminary testing of the new five-level version of EQ-5D (EQ-5D-5L). Quality of life research 2011;20(10):1727-1736. doi: 10.1007/s11136-011-9903-x
[6] MA RJB. After-Sensations and Lingering Pain Following Examination in Patients with Fibromyalgia Syndrome. 2022.
[7] Nicholas MK. The pain self-efficacy questionnaire: taking pain into account. European journal of pain 2007;11(2):153-163. doi: 10.1016/j.ejpain.2005.12.008
[8] Snaith RP. The hospital anxiety and depression scale. Health and quality of life outcomes 2003;1(1):29. doi: 10.1186/1477-7525-1-29
[9] Sullivan MJ, Bishop SR, Pivik J. The pain catastrophizing scale: development and validation. Psychological assessment 1995;7(4):524. doi: 10.1037/1040-3590.7.4.524
[10] Tan G, Jensen MP, Thornby JI, Shanti BF. Validation of the Brief Pain Inventory for chronic nonmalignant pain. The journal of pain 2004;5(2):133-137. doi: 10.1016/j.jpain.2003.12.005
[11] Wolfe F, Clauw DJ, Fitzcharles MA, Goldenberg DL, Katz RS, Mease P, Russell AS, Russell IJ, Winfield JB, Yunus MB. The American College of Rheumatology preliminary diagnostic criteria for fibromyalgia and measurement of symptom severity. Arthritis care & research 2010;62(5):600-610. doi: 10.1002/acr.20140
[12] Wolfe F, Smythe HA, Yunus MB, Bennett RM, Bombardier C, Goldenberg DL, Tugwell P, Campbell SM, Abeles M, Clark P. The American College of Rheumatology 1990 criteria for the classification of fibromyalgia. Arthritis & Rheumatism: Official Journal of the American College of Rheumatology 1990;33(2):160-172. doi: 10.1002/art.1780330203
